# Functional Connectivity Predictors and Mechanisms of Symptom Change in Functional Neurological Disorder

**DOI:** 10.64898/2026.01.27.26344860

**Authors:** Christiana Westlin, Cristina Bleier, Andrew J. Guthrie, Sara A. Finkelstein, Julie Maggio, Jessica Ranford, Julie MacLean, Ellen Godena, Daniel Millstein, Jennifer Freeburn, Caitlin Adams, Christopher D. Stephen, Ibai Diez, David L. Perez

**Affiliations:** Functional Neurological Disorder Unit and Research Group, Mass General Brigham, Massachusetts General Hospital, Harvard Medical School, Boston MA, USA; Mass General Brigham Department of Neurology, Massachusetts General Hospital, Harvard Medical School, Boston, MA, USA; Mass General Brigham Department of Psychiatry, Massachusetts General Hospital, Harvard Medical School, Boston, MA, USA; Athinoula A. Martinos Center for Biomedical Imaging, Massachusetts General Hospital, Harvard Medical School, Boston, MA, USA; Department of Physical Therapy, Massachusetts General Hospital, Boston, MA, USA; Department of Occupational Therapy, Massachusetts General Hospital, Boston, MA, USA; Department of Speech, Language, and Swallowing Disorders, Massachusetts General Hospital, Harvard Medical School, Boston, MA, USA; Computatonal Neuroimaging Lab, Biobizkaia Health Research Institute, Barakaldo, Spain; Ikerbasque Baske Foundation for Science, Bilbao, Spain; Gordon Center for Medical Imaging, Department of Radiology, Massachusetts General Hospital, Harvard Medical School, Boston, MA, USA

**Keywords:** functional movement disorder, functional seizures, fMRI, graph theory, functional connectivity, clinical outcomes

## Abstract

**Background:** Clinical trajectories in functional neurological disorder (FND) are variable, and the mechanisms underlying this heterogeneity remain poorly understood.

**Objective:** This longitudinal study examined resting-state functional connectivity predictors and mechanisms of symptom change in FND.

**Methods:** Thirty-two adults with FND (motor and/or seizure phenotypes) completed baseline questionnaires and a functional MRI (fMRI) session, followed by naturalistic treatment for 6.8±0.8 months. All participants completed follow-up questionnaires; 28 individuals completed a follow-up fMRI. At each timepoint, three graph-theory network metrics of functional connectivity were computed: weighted-degree (centrality), integration (*between-network* connectivity), and segregation (*within-network* connectivity). Analyses adjusted for age, sex, anti-depressants, head motion, time between sessions, and baseline score-of-interest, with cluster-wise correction. Results were contextualized against 50 age-, sex-, and head motion-matched healthy controls (HCs).

**Results:** Based on patient-reported Clinical Global Impression of Improvement, 59.4% improved, 31.3% were unchanged, and 9.3% worsened. Psychometric scores of core FND symptoms and non-core physical symptoms showed variable trajectories, with no group-level changes. Greater improvement in core FND symptoms was associated with higher baseline *between-network* integrated connectivity and reduced integration longitudinally within salience, frontoparietal, and default mode network regions. Right anterior insula integration emerged as a prognostic marker and mechanistic site of reorganization, with the most improved participants showing elevated baseline integration compared to HCs. Increased baseline *within-network* segregated connectivity in dorsal attention network regions correlated with non-core physical symptom improvement. Findings remained significant adjusting for FND phenotype.

**Conclusions:** This study identified large-scale network interactions as potential prognostic and mechanistically-relevant sites of reorganization related to symptom change in FND.

## INTRODUCTION

Functional neurological disorder (FND) is a common, costly, and potentially disabling condition at the neurology-psychiatry intersection, characterized by motor, sensory and cognitive symptoms.^1–3^ Mixed symptoms occur in one-quarter to one-half of individuals, with others developing distinct functional neurological symptoms longitudinally.^4–6^ The most well-studied FND phenotypes are the motor (FND-motor) and seizure (FND-seizure) variants, frequently co-occurring with affective and trauma-related conditions.^5^ The biopsychosocial complexity of FND underscores the importance of multidisciplinary care, with clinical trials and naturalistic cohort studies supporting roles for psychotherapy and physical rehabilitation.^7–10^ Nonetheless, there is individual-level heterogeneity regarding clinical trajectories.^11–14^ As such, there is a need to investigate how brain network organization relates to FND symptom change over time.

Resting-state functional connectivity (rsFC) alterations in somatomotor (SMN), salience (SAN), and default mode (DMN) networks, among others, have been identified across FND cohort studies.^15^ Several studies found aberrant connectivity between the SAN and motor control regions, and between the temporoparietal junction (TPJ) and sensorimotor regions.^16–20^ Work from our group identified altered functional network architecture in FND, marked by increased integration (*between*-*network* connectivity) for SMN regions, with heightened connections to the SAN, DMN, and frontoparietal (FPN) networks.^21^ These findings support the hypothesis that distributed, network-level disruptions play a role in the pathophysiology of FND, with potential relevance as baseline predictors of clinical trajectories and as mechanisms underlying longitudinal symptom change.

Research has begun to explore neuroimaging correlates of clinical trajectories in FND.^17,22–26^ For example, improvements in functional tremor following cognitive behavioral therapy (CBT) were linked to decreased anterior cingulate and paracingulate activations.^23^ Following one-week of multidisciplinary treatment, increased amygdala-ventromedial prefrontal cortex functional connectivity was observed in the subset of FND-motor patients with a favorable response.^24^ Improved outcomes in a mixed FND cohort have been associated with greater supplementary motor area signal variability, whereas worse outcomes were associated with higher baseline left insula variability.^25^ In earlier FND research, we identified an association between improved symptom scores and relative baseline increases in left centromedial amygdala-to-right anterior insula rsFC.^17^ Nonetheless, much of the literature has relied on region-of-interest (ROI) approaches, examined either baseline predictors or post-treatment mechanisms in isolation, or focused on narrowly defined subpopulations with limited real-world relevance (e.g., euthymic individuals only^22^). This underscores the need for longitudinal, whole-brain investigations that examine both predictors and mechanisms of symptom change in a representative FND cohort.

Here, we employed rsFC graph-theory analyses to examine baseline predictors (*n=*32) and longitudinal mechanisms (*n=*28) of symptom change in FND. Patients underwent baseline and six-month follow-up neuroimaging, between which they received individualized, naturalistic treatment as part of usual care. At both timepoints, we assessed three network organization metrics: weighted-degree (centrality), integration (*between-network* connectivity), and segregation (*within-network* connectivity). Our goal was to identify baseline network features, and longitudinal changes in network organization, that related to individual differences in symptom change. By linking brain architecture to clinical trajectories, this work aims to advance FND mechanistic models and guide future development of personalized, neurobiologically-informed interventions.

## METHODS

### Participants

Thirty-two consecutive participants with FND (28 females, 4 males; mean age=42.4±13.3; average illness duration 3.7±4.2 years (range=0.1-16.1 years)) were recruited from the Massachusetts General Hospital FND Unit between March 2021 and October 2024 (**Table 1**). FND diagnoses were based on positive examination signs, semiological features, and electroencephalography data (FND-seizure only^27^).^28^ Participants with either FND-motor, FND-seizure, or both were eligible, reflecting observations that individuals often present with mixed symptoms and/or develop new FND symptoms over time.^5^ Exclusion criteria included major neurological comorbidities (e.g., epilepsy, Parkinson’s disease), known brain MRI abnormalities, poorly controlled medical conditions with central nervous system (CNS) effects, active illicit substance dependence, psychosis, and/or active suicidality. Twenty-seven had FND-motor (tremor=12; gait=12; speech=12; weakness=8; tics/jerks/spasms=5; dystonia=2; presentations were not mutually exclusive) and 10 had FND-seizure (documented=6; probable=4); 5 participants had FND-motor and FND-seizure (see **Supplementary Fig. 1** for a depiction of FND symptoms across participants). Only cross-sectional [baseline] neuroimaging data for this cohort has been previously published.^21,29,30^ Twenty-eight of 32 individuals fully completed the follow-up session (four provided follow-up questionnaire data only; see **Supplementary Table 1** sub-cohort demographics). One participant changed SSRI/SNRI use status during the study period and was included only in predictor analyses; SSRI/SNRI use remained stable for all participants included in longitudinal analyses.

**Table 1.** Clinical characteristics of the longitudinal functional neurological disorder cohort.

| | **Baseline**<br>**(n=32)**<br>mean ± SD or N (%) | **Follow-Up**<br>**(n=32)**<br>mean ± SD or N (%) | **Uncorrected**<br>**p-value<sup>a</sup>** |
| --- | --- | --- | --- |
| **Demographics** |  |  |  |
| Age at consent (yrs.) | 42.4 ± 13.3; range: 22.4-72.1;<br>median: 44.0 | - | - |
| Biological sex | 28F; 4M | - | - |
| Illness duration (yrs.) | 3.7 ± 4.2; range: 0.1-16.1;<br>median: 2.0 | - | - |
| Interval follow-up period<br>(months) | 6.8 ± 0.8; range 5.4-8.8; median<br>6.6 | - | - |
| Race (white) | 28 (87.5%) | - | - |
| Married | 15 (46.9%) | 15 (46.9%) | 1.00 |
| College graduate | 22 (68.8%) | - | - |
| Employed full-time (or full-time student) | 12 (37.5%) | 14 (43.8%) | 0.69 |
| On medical disability or<br>worker's compensation | 10 (31.3%) | 6 (18.8%) | 0.25 |
| Functional motor disorder | 27 (84.4%) | - | - |
| Functional seizures | 10 (31.3%) | - | - |
| SSRI/SNRI use | 13 (40.6%) | 14 (43.8%) | 1.00 |
| CGI-I | - | Improved<sup>b</sup>: 19 (59.4%)<br>No change: 10 (31.3%)<br>Worsened: 3 (9.3%) | - |
| **Psychometric Scores** |  |  |  |
| SOMS:CD | 6.6 ± 6.3; range: 0-21; median:<br>5 | 5.7 ± 5.8; range: 0-22; median:<br>4.5 | 0.21 |
| PHQ-15 | 11.8 ± 5.7; range: 1-26; median:<br>11.0 | 12.2 ± 5.7; range: 3-25; median:<br>12.0 | 0.55 |
| BDI-II | 15.3 ± 11.4; range: 0-38;<br>median: 13.0 | 12.1 ± 11.3; range 0-40; median:<br>9.5 | 0.018 |
| STAI-total | 82.5 ± 23.2; range 49-140;<br>median: 77.5 | 77.3 ± 21.2; range 44-120;<br>median 78.5 | 0.038 |
| PCL-5 | 23.4 ± 17.9; range: 0-62;<br>median: 20 | - | - |
| CTQ-abuse | 31.0 ± 14.7; range: 15-68;<br>median: 27.5 | - | - |
| CTQ-neglect | 21.6 ± 8.5; range: 10-37;<br>median: 19.5 | - | - |

<sup>a</sup> p-value corresponds to uncorrected statistic for baseline vs follow-up comparisons. For psychometric scores, FDR-corrected p-values are as follows: SOMS:CD: 0.28, PHQ-15: 0.55, BDI-II: 0.072, STAI-total: 0.076. For demographic variables, McNemar tests were used to assess married, employed, medical disability, and SSRI/SNRI use variables; for psychometric score variables, paired samples t-tests were used to assess pre-post changes for PHQ-15 and STAI-total scores (normally distributed variables), and Wilcoxon signed-rank tests were used for SOMS:CD and BDI-II (non-normally distributed). <sup>b</sup>Improved includes: "much improved" (n=5) and "improved" (n=14) based on patient-reported Clinical Global Impression of Improvement (CGI-I) scale scores; SOMS:CD, Screening for Somatoform Symptoms-7 Subscale for Conversion Disorder; PHQ-15, Patient Health Questionnaire-15; BDI-II, Beck Depression Inventory-II; STAI, Spielberg State-Trait Anxiety Inventory; PCL-5, PTSD Checklist 5; CTQ, Childhood Trauma Questionnaire.

Five additional participants were excluded following imaging acquisition and processing due to excessive head motion (n=4) or following outlier inspection based on mean weighted-degree (n=1) as previously described.^21^ Twenty additional participants provided baseline psychometric and imaging data but did not complete at least the follow-up questionnaires (**Supplementary Table 2**). These participants were classified as lost to research follow-up if they did not complete the follow-up session after at least two contact attempts.

Fifty healthy controls (HCs, 43 females, 7 males; mean age=38.7±10.5 years; mean head motion [framewise displacement]=0.06±0.03) were also included for *post-hoc* comparisons (**Supplementary Table 3**). These participants were prospectively recruited from the community and had no psychiatric, major neurological, or poorly controlled medical conditions with CNS effects. HCs were matched to the FND cohort based on age, sex, and head motion. All subjects signed informed consent, and the Mass General Brigham Institutional Review Board approved this study.

### Study Design and Individualized, Naturalistic Treatment

At baseline, participants had an MRI scan, underwent a Structured Clinical Interview (SCID-DSM-5), and completed self-report questionnaires. Approximately six months later (mean 6.8±0.8 months), participants returned for a repeat MRI scan and follow-up questionnaires.

Between baseline and follow-up sessions, participants received naturalistic, individualized treatment [usual care at our institution] as determined collaboratively by the patient and FND Unit team members. All participants remained in longitudinal care with an FND Unit neurologist or neuropsychiatrist. As increasingly endorsed^31^, treatment recommendations were individualized based on: i) phenotype, ii) concurrent neuropsychiatric and psychosocial factors^7^, iii) published recommendations^32–34^, iv) original research protocols^8,35^ – including treatment studies from our own group^36,37^, v) patient preferences, and vi) travel and insurance considerations among other factors. Interventions included psychotherapy, physiotherapy, occupational therapy, and/or speech and language therapy. Treatment was not limited to our healthcare system, and thus a subset received care across our academic medical center and the community.

### Neuropsychiatric Characterization and Longitudinal Monitoring

At baseline, participants completed questionnaires to assess symptom severity and other neuropsychiatric features. Core FND symptom burden was evaluated using the Screening for Somatoform Symptoms-7 Subscale for Conversion Disorder (SOMS:CD), a questionnaire of FND symptoms (e.g., paralysis, impaired balance, seizures) experienced during the past week, rated on a 5-point Likert scale.^38^ Non-core physical symptom severity was assessed with the Patient Health Questionnaire-15 (PHQ-15), which captures bothersome physical symptoms (e.g., pain, fatigue) during the previous four weeks, using a 3-point Likert scale.^39^ Participants also completed the Beck Depression Inventory-II (BDI-II), Spielberger State-Trait Anxiety Inventory (STAI), PTSD Checklist-5 (PCL-5), and the Childhood Trauma Questionnaire (CTQ). At follow-up, participants completed the SOMS:CD, PHQ-15, BDI-II and STAI, along with laboratory-designed questions to assess the treatments received in the interval period (see **Supplementary Methods**); participants also completed the Clinical Global Impression of Improvement scale (CGI-I) to evaluate their perceived change in FND clinical status, based on a 5-point Likert scale (“much improved” to “much worse”).

### MRI Acquisition and Preprocessing

3T MRI acquisition and preprocessing details are described in **Supplementary Materials.** Anatomical and functional MRI data were preprocessed using FMRIB Software Library v5.0.7 and MATLAB 2023a using preprocessing pipelines as described previously.^21,40^

### Resting-State Functional Connectivity

Voxel-wise network properties were quantified using three graph theory metrics - weighted-degree, integration, and segregation - as previously described^21^; see **Fig. 1** for an overview. rsFC matrices were computed per participant by calculating Pearson correlation coefficients between the time series of each pair of grey matter voxels. Negative correlations were removed due to their controversial interpretation.

**Figure 1.**
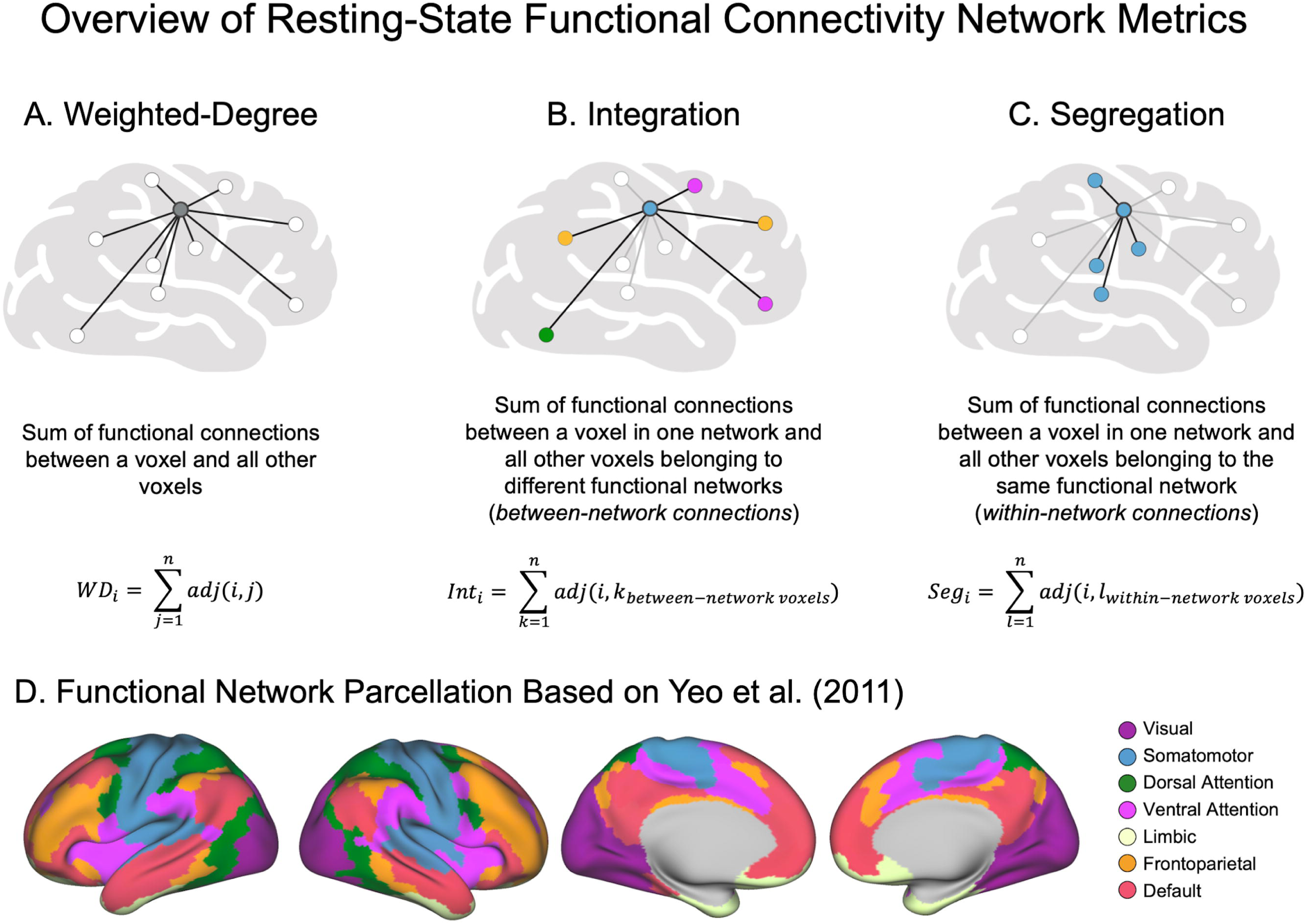
Overview of graph theory-based resting-state functional connectivity metrics used in the present study. (**A**) Weighted-degree is computed by summing the weighted connections for a given voxel and all other voxels. A higher weighted-degree therefore indicates that a given voxel is more globally connected to the rest of the brain. (**B**) Integration is computed by summing the isocortical weighted connections for a given voxel in one network and all other voxels belonging to a *different* functional network than the originating voxel. A high integration value therefore indicates that a given voxel is important for between-network communication. (**C**) Segregation is computed by summing the isocortical weighted connections for a given voxel in one network and all other voxels belonging to the *same* network. A high segregation value therefore indicates that a given voxel is important for within-network modular communication. For each of these metrics, only positive connections were summed. (**D**) Visualization of the Yeo et al. (2011) seven-network parcellation used to assign voxels to resting-state networks in integration and segregation analyses.

Weighted-degree was calculated from whole-brain rsFC matrices to assess the global connectivity of each voxel. For a given voxel *i*, weighted-degree (WD) was defined as the sum of its weighted positive connections:

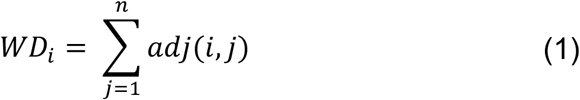

where *adj(i,j)* is the weighted functional connection between voxels *i* and *j*. The result was a subject-level map indicating how strongly each voxel was connected to the rest of the brain.

Integration and segregation metrics were computed from isocortical-only rsFC matrices, in which each voxel was assigned to one of seven functional networks based on Yeo et al. (2011): SMN, dorsal attention (DAN), ventral attention (referred to here as salience [SAN]), FPN, DMN, limbic (LIM) and visual (VIS) networks.^41^ Connections between voxels in *different* networks were categorized as integration connections, whereas connections within the *same* network were labeled as segregation connections. For each voxel *i*, integration and segregation were defined as:

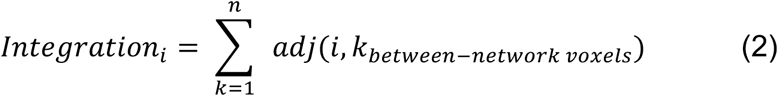

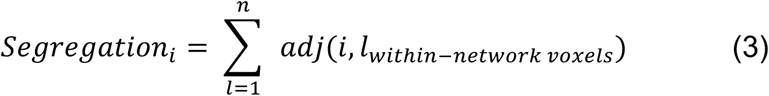

These calculations each yielded a voxel-wise map per participant, with the integration map reflecting the extent to which each voxel integrated across networks, and the segregation map reflecting its within-network connectivity.

### Statistical Analyses

#### Baseline rsFC Predictors of Symptom Change

Symptom change was computed as the difference between follow-up and baseline scores for core FND symptoms (SOMS:CD), and separately for non-core physical symptoms (PHQ-15). Separate models were run for each rsFC measure (weighted-degree, integration, segregation) and symptom change scores. All general linear models (GLMs) controlled for age, sex, SSRI/SNRI use (yes/no), mean FD, time between baseline and follow-up, and the baseline symptom score of interest (SOMS:CD or PHQ-15). Findings were corrected for multiple comparisons using cluster-wise correction via Monte Carlo simulation with 10,000 iterations, estimating the probability of observing false positive clusters at *p*<0.05. *Post-hoc* analyses were conducted to assess whether findings remained significant adjusting for: (i) FND phenotype (FND-seizure yes/no); (ii) baseline BDI-II, STAI-total, and PCL-5 scores; and (iii) CTQ-abuse and CTQ-neglect scores.

#### Longitudinal Changes in rsFC Associated with Symptom Change

To examine whether longitudinal changes in rsFC network organization were associated with clinical trajectories, we conducted the same series of voxel-wise GLMs described in the above baseline predictors section, using updated independent variables. Specifically, the independent variables were rsFC change maps for each network measure (weighted-degree, integration, segregation), computed as the difference between follow-up and baseline rsFC maps. These GLM analyses included the same primary and *post-hoc* adjustments as described above, with two additional *post-hoc* adjustments: (i) change in BDI-II scores, and (ii) change in STAI-total scores. No participants in this analysis had a change in SSRI/SNRI use (yes/no). Participants did not differ based on mean FD between baseline (0.073±0.041) and follow-up (0.076±0.032), *t*(27)=0.41, *p*=0.68.

### *Post-Hoc* Connectivity Analyses to Characterize Integration Findings

To understand which functional networks the voxels with significant integration-symptom change associations were integrating across, we conducted *post-hoc* seed-based connectivity analyses. Specifically, we assigned each voxel that showed a significant association between integration-related rsFC and symptom change to one of the seven Yeo et al. (2011) networks, and created a seed ROI for each network by averaging the time series across all voxels within that network. For the top three networks containing the most significant voxels, we then computed Pearson correlations between the average time series of the seed and the time series of all isocortical voxels, and transformed these values using Fisher’s r-to-z transformation. The resulting subject-level connectivity maps were entered into voxel-wise GLMs, testing whether rsFC values significantly differed from zero while adjusting for age, sex, SSRI/SNRI use, and mean FD. Results were corrected for multiple comparisons using cluster-wise correction via Monte Carlo simulation with 10,000 iterations to estimate the probability of false positive clusters with *p*<0.05.

To quantify the network-level connectivity patterns, each significant voxel in the resulting connectivity maps was labeled according to the Yeo et al. (2011) seven-network parcellation. We then computed the number of significant connections from each seed network to each of the six other functional networks (excluding within-network links that are not relevant for measuring integration).

### *Post-Hoc* Contextualization with HCs

To further contextualize findings, we conducted an exploratory comparison with HCs. We identified overlapping voxels that emerged in both baseline predictor and longitudinal change analyses, and extracted mean values across these voxels in HCs and in FND participants at baseline and follow-up. Mann-Whitney U tests were used to compare mean values of HCs vs. FND participants, across the full FND cohort and stratified by the top/bottom 20% of individuals who reported the greatest symptom improvement/worsening.

## RESULTS

### Clinical Observations

Among the 32 participants, 30 (94%) received psychotherapy, with 21 (65.6%) specifically reporting receipt of CBT. Seventeen participants (53%) reported receiving physiotherapy, 16 (50%) endorsed receiving occupational therapy, and 10 (31%) reported receiving speech and language therapy. Eleven participants (34.4%) had all FND care within our healthcare system; the remainder either received a combination of treatment within our healthcare system and the community (n=16, 50.0%) or entirely in the community (n=5, 15.6%), limiting our ability to objectively capture treatment-related frequency and duration information (**Table 1** and **Supplementary Table 4**).

Based on CGI-I scores, 5 participants (15.6%) reported being “much improved”, 14 (43.8%) “improved”, 10 (31.3%) “unchanged”, and 3 (9.3%) “worse”; no participants rated themselves as “much worse”. SOMS:CD and PHQ-15 scores showed no group-level differences at follow-up vs. baseline (Wilcoxon signed-rank tests: SOMS:CD: *z*=-1.25, *p*=0.21; PHQ-15: *z*=-0.43, *p*=0.67). However, scores showed prominent individual differences. For SOMS:CD scores, 14 participants reported reduced symptoms (mean change=-4.4±3.3; median=-3), 9 reported no change, and 9 endorsed increased symptoms (mean change=3.9±4.3; median=3). For PHQ-15 scores, 15 participants reported reduced symptoms (mean change=-2.6±1.2; median=-3), 1 reported no change, and 16 endorsed increased symptoms (mean change=3.1±1.8; median=3) (**Table 1**).

No baseline demographic or psychometric variables were associated with symptom improvement (SOMS:CD change scores; PHQ-15 change scores; CGI-I improved vs. not-improved) following false discovery rate correction at the level of each symptom measure. However, at an uncorrected threshold (p<0.05), those who reported improvement on the CGI-I had shorter illness durations (3.2±4.7 years vs. 4.5±3.4 years) and were less likely to have FND-seizure (**Supplementary Table 5**). No baseline characteristics showed uncorrected associations with SOMS:CD or PHQ-15 change scores.

Compared to longitudinal cohort participants (n=32), those lost to follow-up (n=20) showed no significant between-group differences on available baseline characteristics (**Supplementary Table 2**).

### Baseline Functional Connectivity Predictors of Symptom Change

#### Weighted-Degree

Baseline rsFC weighted-degree values in the right middle frontal and precentral gyri, and left cerebellum, were positively associated with changes in SOMS:CD scores at follow-up compared to baseline (**Fig. 2A**). Specifically, higher baseline weighted-degree in these areas was linked to greater FND symptom improvement, while lower baseline weighted-degree in these regions related to symptom worsening. All findings held adjusting *post-hoc* for phenotype (FND-seizure yes/no), but only the cerebellar findings remained significant for CTQ-abuse and CTQ-neglect scores; no findings held adjusting for baseline BDI-II, STAI-total or PCL-5 scores (**Supplementary Fig. 2**). No regions showed an association between baseline weighted-degree values and PHQ-15 score changes.

**Figure 2.**
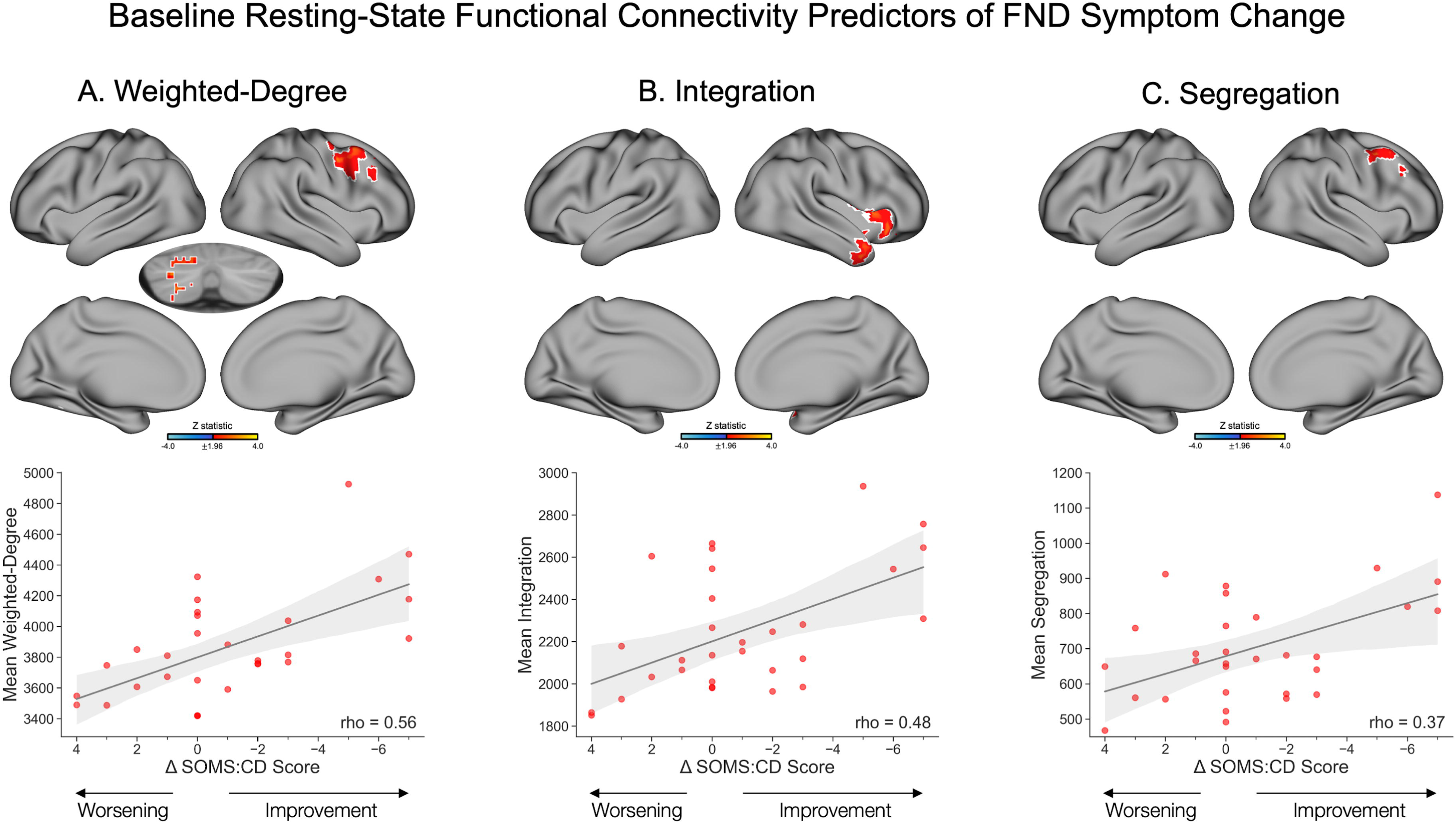
Baseline resting-state functional connectivity predictors of functional neurological disorder (FND) symptom change, as measured by a change in SOMS:CD scores between baseline and follow-up. (**A**) *Top panel* depicts regions with baseline weighted-degree values that were significantly associated with FND symptom change. Specifically, baseline weighted-degree in the right middle frontal gyrus, right precentral gyrus, and left cerebellum was positively associated with FND symptom change, such that a higher baseline weighted-degree in these regions was associated with greater symptom improvement, and a lower weighted-degree was associated with a greater worsening of symptoms. *Bottom panel* depicts a scatterplot of SOMS:CD change scores on the x-axis and mean weighted-degree values averaged across significant voxels on the y-axis. (**B**) *Top panel* depicts a positive association between baseline integration values in the right insula and right temporal pole and FND symptom change scores. Increased baseline integration in these regions was associated with symptom improvement, while decreased baseline integration was associated with symptom worsening. *Bottom panel* depicts a scatterplot of SOMS:CD change scores on the x-axis and mean integration values averaged across significant voxels on the y-axis. (**C**) *Top panel* depicts a positive association between baseline segregation values in the right middle frontal precentral gyri and SOMS:CD change scores, such that increased segregation in these regions was associated with symptom improvement, and decreased segregation was associated with symptom worsening. *Bottom panel* depicts a scatterplot of SOMS:CD change scores on the x-axis and mean segregation values averaged across significant voxels on the y-axis. Volumetric brain maps are visualized on the surface for display purposes only; subcortical results in volume are shown with the left hemisphere on the left side of the image. Colors in the maps reflect the z-statistic for the primary general linear model adjusting for age, sex, SSRI/SNRI use, mean framewise displacement, elapsed time between baseline and follow-up sessions, and baseline SOMS:CD score (z-statistic>1.96; p<0.05 cluster-corrected for multiple comparisons). White outlines reflect regions that also held across *post-hoc* corrections for FND phenotype (FND-seizure yes/no). Statistical maps for additional *post-hoc* corrections are visualized in **Supplementary Fig. 2**. While analyses were conducted at the voxel-wise level, Spearman correlation coefficients are included in the scatterplots to illustrate the strength of the associations. Shaded regions reflect 95% confidence intervals. For visualization purposes, change scores are plotted with negative values on the right to reflect symptom improvement. Additionally, two participants with SOMS:CD change scores that exceeded more than 1.5 times the interquartile range above the upper quartile or below the lower quartile were excluded from the plots for clarity, but retained in analyses. SOMS:CD, Screening for Somatoform Symptoms-7 Subscale for Conversion Disorder.

#### Integration

Baseline integration values in the right anterior insula and temporal pole were positively associated with changes in SOMS:CD scores. Specifically, higher baseline integration in these regions related to longitudinal FND symptom improvement, and lower baseline integration values related to symptom worsening (**Fig. 2B**). These findings held when adjusting *post-hoc* for phenotype (FND-seizure yes/no) but did not hold when adjusting for baseline affective symptoms or childhood trauma (**Supplementary Fig. 2**). No regions showed an association between baseline integration values and PHQ-15 score changes.

#### Post-hoc Characterization of Baseline Integration-Based Network Connectivity

To investigate what networks were connected to the regions that showed associations between baseline integration and SOMS:CD change scores, we conducted *post-hoc* seed-to-voxel connectivity analyses. Most of the identified voxels were within SAN, FPN, and DMN. The SAN seed at baseline showed increased rsFC largely to SMN, FPN, and DMN regions, while the FPN seed showed increased connectivity largely to DMN, SAN, and SMN regions; the DMN seed showed increased connectivity largely to LIM, FPN, SAN, and SMN (**Fig 3., Supplementary Fig. 3**).

**Figure 3.**
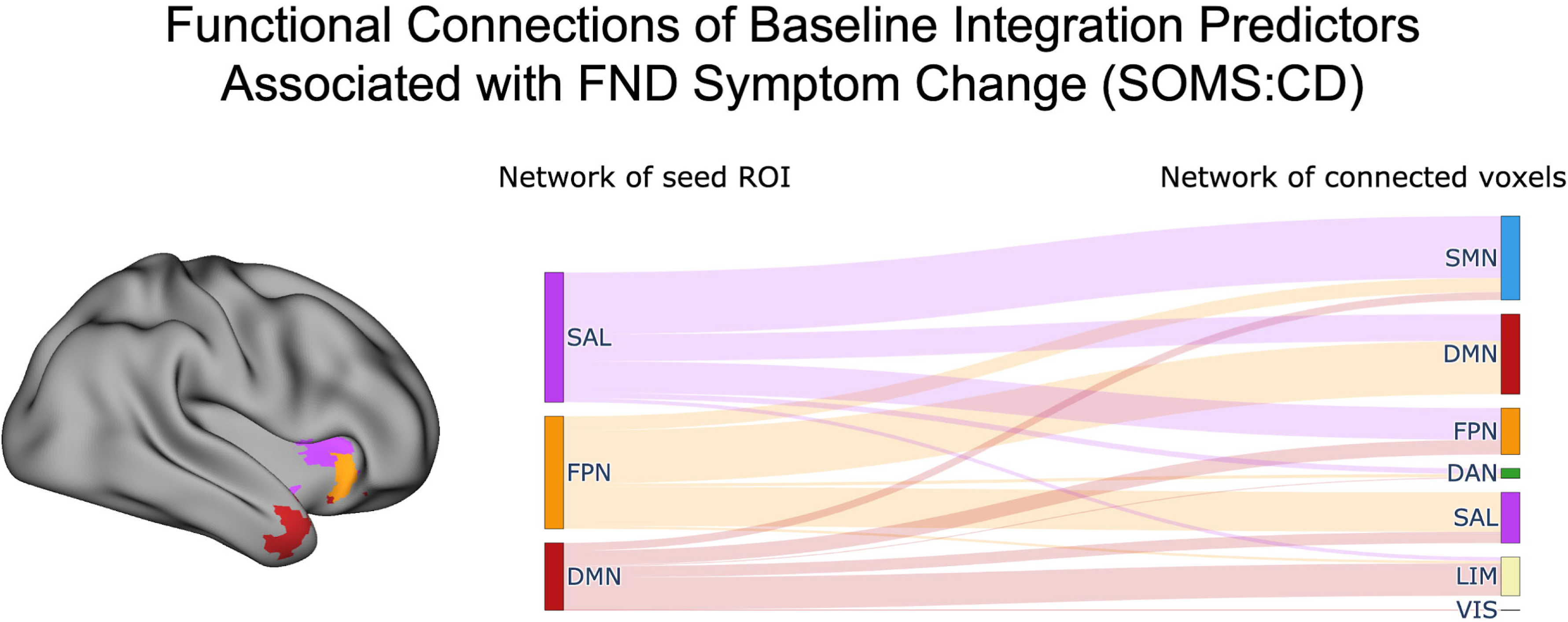
Network connectivity of baseline integration predictors associated with functional neurological disorder (FND) symptom change. *Left panel* shows voxels whose baseline integration values were significantly associated with FND symptom change (Δ SOMS:CD scores), colored according to their functional network assignment based on the Yeo et al. (2011) 7-network parcellation. Only the three networks containing the most significant voxels are displayed: salience (SAN; magenta), frontoparietal (FPN; orange), and default mode network (DMN; red). For each of these networks, we extracted the average time series of all significant voxels for use in seed-to-voxel connectivity analyses, to then characterize the networks they were functionally integrating across. *Right panel* depicts a Sankey diagram summarizing the between-network connectivity patterns for each seed. The left side of the diagram denotes the seed network, while the right side indicates the network identity of significantly connected voxels. Line thickness reflects the number of significant connections, with thicker lines indicating a higher number of between-network connections. Self-connections (e.g., DMN-to-DMN) were excluded, as integration analyses focused on between-network connectivity. See **Supplementary Figure 3** for detailed connectivity breakdowns across all networks. SMN, somatomotor network; DMN, default mode network; FPN, frontoparietal network; DAN, dorsal attention network; SAN, salience network; LIM, limbic network; VIS, visual network; SOMS:CD, Screening for Somatoform Symptoms-7 Subscale for Conversion Disorder.

#### Segregation

Baseline segregation values in FPN regions of the right middle frontal and precentral gyri were positively associated with a longitudinal change in SOMS:CD scores, such that symptom improvement was linked to higher baseline segregation connections, and symptom worsening was linked to lower baseline segregation connections (**Fig. 2C**). These findings held when adjusting *post-hoc* for phenotype (FND-seizure yes/no), and partially held when adjusting for baseline BDI-II, STAI-total, and PCL-5 scores; findings did not hold when adjusting for CTQ-abuse and CTQ-neglect scores (**Supplementary Fig. 2**).

Baseline segregation values in the left post-central and supramarginal gyri of the DAN were associated with a change in PHQ-15 scores, with an increase in baseline segregation linked to physical symptom improvement at follow-up, and a decrease in baseline segregation linked to physical symptom worsening (**Fig. 4**). Findings held across *post-hoc* adjustments for (i) phenotype (FND-seizure yes/no) and (ii) baseline BDI-II, STAI-total, and PCL-5 scores; findings did not remain significant adjusting *post-hoc* for CTQ-abuse and CTQ-neglect scores (**Supplementary Fig. 4**).

**Figure 4.**
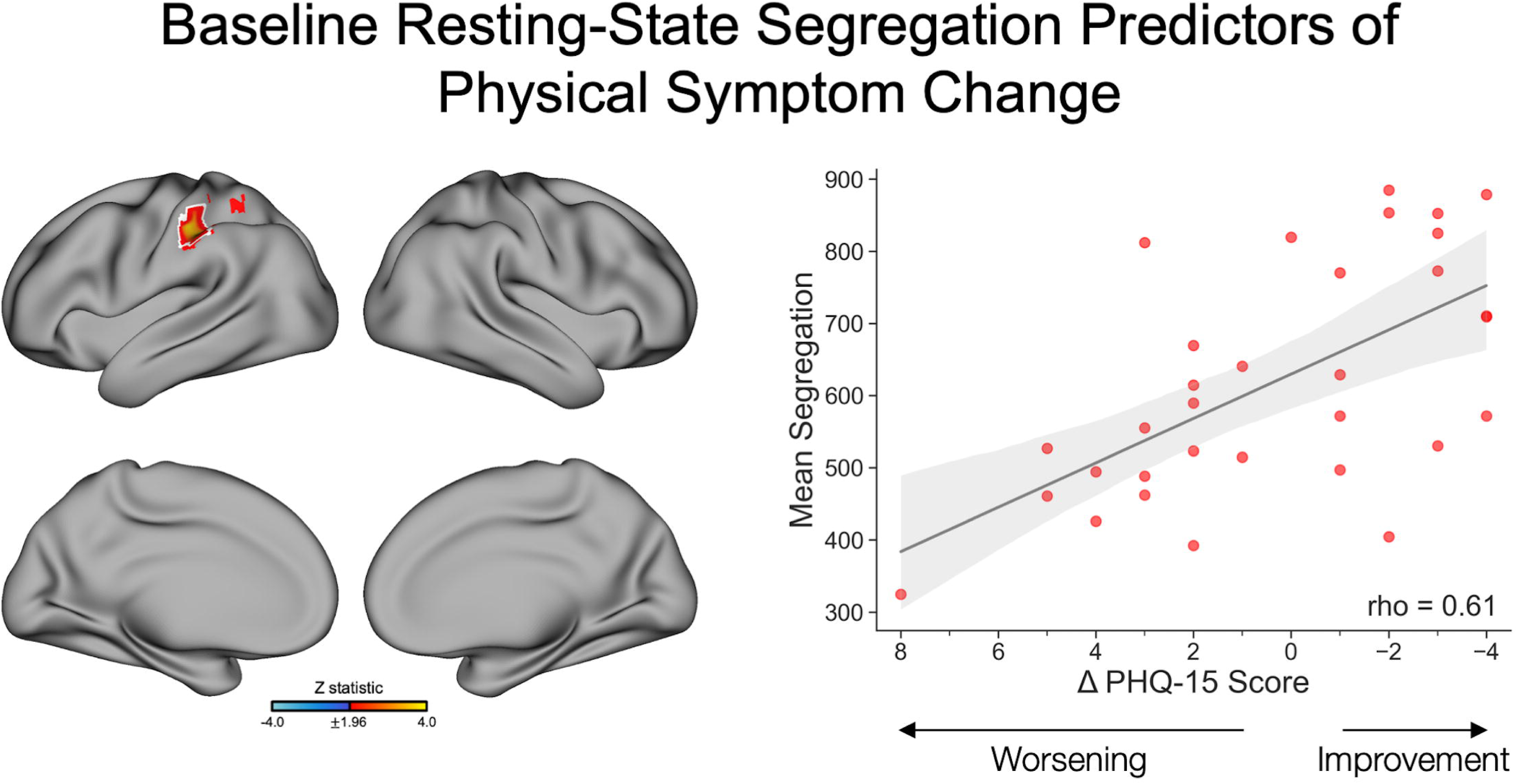
Baseline resting-state segregation predictors of physical symptom change, as measured by a change in PHQ-15 scores between baseline and follow-up. *Left panel* depicts regions with baseline segregation values that were significantly associated with changes in self-reported physical symptoms. Specifically, baseline segregation in the left post-central and left supramarginal gyri was positively associated with physical symptom change, such that a higher baseline segregation in these regions was associated with greater symptom improvement, and a lower segregation was associated with a greater worsening of symptoms. The volumetric brain map is visualized on the surface for display purposes only. Colors in the map reflect the z-statistic for the primary general linear model adjusting for age, sex, SSRI/SNRI use, mean framewise displacement, elapsed time between baseline and follow-up sessions, and baseline PHQ-15 score (z-statistic>1.96; p<0.05 cluster-corrected for multiple comparisons). White outlines reflect regions that also held across *post-hoc* correction for FND phenotype (FND-seizure yes/no). Statistical maps for additional *post-hoc* corrections are visualized in **Supplementary Fig. 4**. *Right panel* depicts a scatterplot of PHQ-15 change scores on the x-axis and mean segregation values averaged across significant voxels on the y-axis. While the analysis was conducted at the voxel-wise level, a Spearman correlation coefficient is included in the scatterplots to illustrate the strength of the association. The shaded region reflects the 95% confidence interval. For visualization purposes, change scores are plotted with negative values on the right to reflect symptom improvement. PHQ-15, Patient Health Questionnaire-15.

### Longitudinal Changes in Functional Connectivity Associated with Symptom Change

#### Weighted-Degree

Longitudinal rsFC weighted-degree changes in the right precentral gyrus, superior parietal lobule, lateral occipital cortex, and cerebellum were negatively associated with SOMS:CD symptom changes between follow-up and baseline (**Fig. 5A**). Specifically, as weighted-degree values decreased at follow-up compared to baseline, symptom scores improved. Cerebellar findings held across all *post-hoc* adjustments. Cortical findings held across *post-hoc* adjustments for (i) phenotype (FND- seizure yes/no); (ii) CTQ-abuse and CTQ-neglect scores; (iii) BDI-II score change; and (iv) STAI-total score change but did not remain significant when adjusting for baseline BDI-II, STAI-total, and PCL-5 scores (**Supplementary Fig. 5**). No regions showed an association between weighted-degree changes and PHQ-15 score changes.

**Figure 5.**
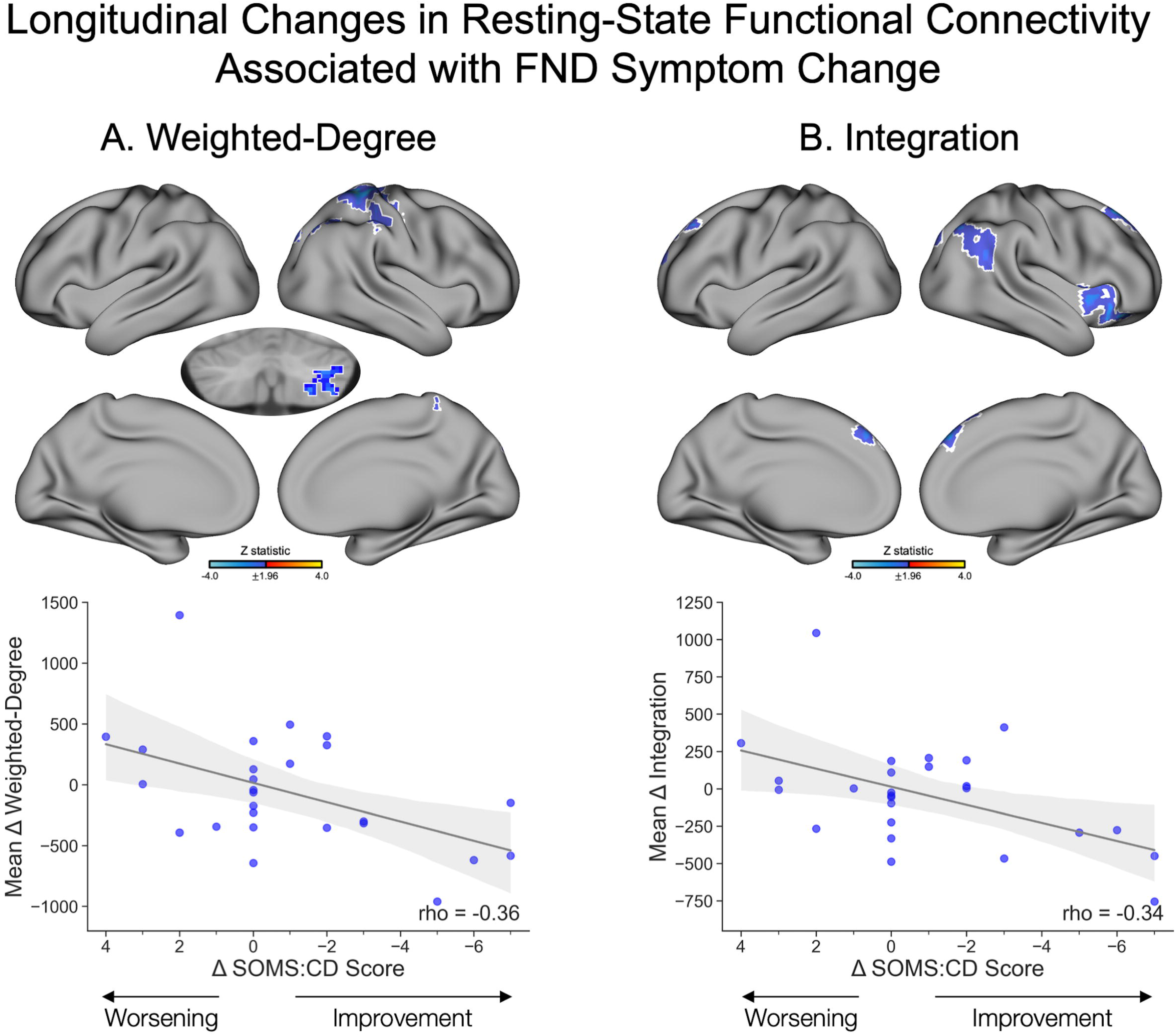
Longitudinal changes in resting-state functional connectivity associated with functional neurological disorder (FND) symptom change (Δ SOMS:CD scores) between baseline and follow-up. (**A**) *Top panel* depicts regions with changes in weighted-degree that were significantly negatively associated with FND symptom change. Specifically, symptom improvement was associated with decreased weighted-degree at follow-up compared to baseline in the right lateral occipital cortex, superior parietal lobule, precentral gyrus, and cerebellum, while increased weighted-degree in these regions at follow-up compared to baseline was associated with symptom worsening. *Bottom panel* depicts a scatterplot of SOMS:CD change scores on the x-axis and mean weighted-degree change values averaged across significant voxels on the y-axis. (**B**) *Top panel* depicts a negative association between integration change and FND symptom change between baseline and follow-up. Symptom improvement was associated with decreased integration at follow-up compared to baseline in the right anterior insula and orbitofrontal cortex, angular gyrus, lateral occipital cortex, and bilateral frontal pole and superior frontal gyrus, and symptom worsening was associated with increased integration at follow-up compared to baseline in these regions. *Bottom panel* depicts a scatterplot of SOMS:CD change scores on the x-axis and mean integration change values averaged across significant voxels on the y-axis. Volumetric brain maps are visualized on the surface for display purposes only; subcortical results in volume are shown with the left hemisphere on the left side of the image. Colors in the maps reflect the z-statistic for the primary general linear model adjusting for age, sex, SSRI/SNRI use, mean framewise displacement, elapsed time between baseline and follow-up sessions, and baseline SOMS:CD score (z-statistic>1.96; p<0.05 cluster-corrected for multiple comparisons). White outlines reflect regions that also held across *post-hoc* corrections for FND phenotype (FND-seizure yes/no). Statistical maps for additional *post-hoc* corrections are visualized in **Supplementary Fig. 5**. While analyses were conducted at the voxel-wise level, Spearman correlation coefficients are included in the scatterplots to illustrate the strength of the associations. Shaded regions reflect 95% confidence intervals. For visualization purposes, change scores are plotted with negative values on the right to reflect symptom improvement. Additionally, two participants with SOMS:CD change scores that exceeded more than 1.5 times the interquartile range above the upper quartile or below the lower quartile were excluded from the plots for clarity, but retained in analyses. SOMS:CD, Screening for Somatoform Symptoms-7 Subscale for Conversion Disorder.

#### Integration

Longitudinal integration changes were negatively associated with SOMS:CD score changes in the right anterior insula, orbitofrontal cortex, angular gyrus, lateral occipital cortex, and bilateral frontal pole and superior frontal gyri (**Fig. 5B**). Specifically, as integration in these regions decreased at follow-up compared to baseline, symptom scores improved. Findings largely held across all *post-hoc* corrections aside from baseline BDI-II, STAI-total, and PCL-5 adjustments; additionally, the right insula finding did not hold when adjusting for CTQ-abuse and CTQ-neglect, or STAI-total change scores (**Supplementary Fig. 5**).

Longitudinal integration changes in the left lateral occipital pole and occipital fusiform gyrus were positively associated with PHQ-15 score changes (**Supplementary Fig. 6**), such that increased integration at follow-up compared to baseline was linked to physical symptom improvement. These findings did not hold for *post-hoc* FND phenotype or CTQ adjustments but held for all other corrections (**Supplementary Fig. 7**).

#### Post-hoc Characterization of Longitudinal Integration-Based Network Connectivity Changes

We again investigated which networks were functionally connected to the regions that showed significant associations between integration changes [baseline vs. follow-up rsFC] and SOMS:CD score changes via *post-hoc* seed-to-voxel connectivity analyses. Most identified voxels were within SAN, FPN, and DMN, which were then used as seeds in this analysis. Compared to baseline rsFC observations, the SAN seed showed decreased connectivity at follow-up to SMN and DMN; the FPN seed showed decreased connectivity at follow-up to DAN, LIM, SAN, and SMN; the DMN seed showed decreased connectivity at follow-up to LIM and FPN (**Fig. 6**).

**Figure 6.**
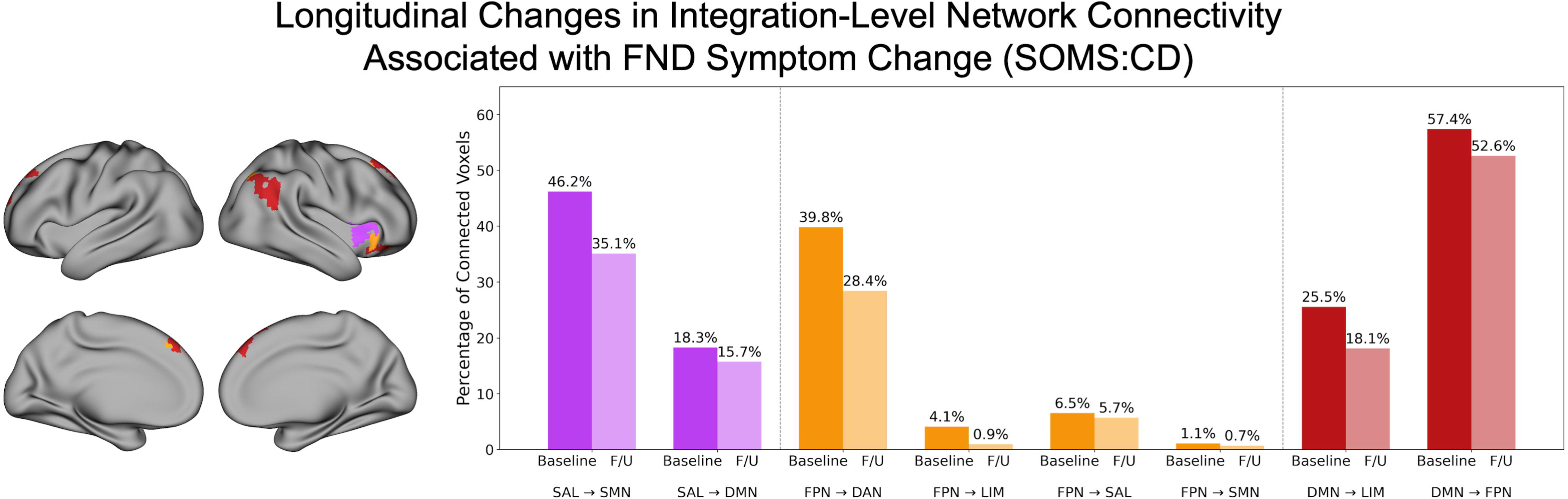
Longitudinal changes in integration-level network connectivity associated with functional neurological disorder (FND) symptom change. *Left panel* shows voxels whose change in integration from baseline to follow-up was significantly associated with FND symptom change (Δ SOMS:CD scores), colored according to their functional network assignment based on the Yeo et al. (2011) 7-network parcellation. Only the three networks containing the most significant voxels are displayed: salience (SAN; magenta), frontoparietal (FPN; orange), and default mode network (DMN; red). For each of these networks, we extracted the average time series of all significant voxels for use in seed-to-voxel connectivity analyses, to then characterize the networks they were functionally integrating across. *Right panel* depicts a bar chart summarizing the between-network connectivity decreases for each seed. Darker bars reflect the percent of connected voxels at baseline, while lighter bars reflect the percent of connected voxels at follow-up. Only networks with decreased connections at follow-up compared to baseline are shown, given that a decrease in integration was associated with symptom improvement. For the SAN seed, connections with the SMN and DMN were relatively lower at follow-up compared to baseline. For the FPN seed, connections with LIM, SAN, and SMN were relatively lower at follow-up compared to baseline. For the DMN seed, connections with LIM and FPN were relatively lower at follow-up compared to baseline. F/U, follow-up; SMN, somatomotor network; DMN, default mode network; FPN, frontoparietal network; DAN, dorsal attention network; SAN, salience network; LIM, limbic network. SOMS:CD, Screening for Somatoform Symptoms-7 Subscale for Conversion Disorder.

We also investigated which networks were functionally connected to the regions that showed significant associations between integration changes and PHQ-15 score changes. Significant voxels fell within the VIS network, and showed increased connectivity with FPN, LIM, and DMN regions at follow-up compared to baseline (**Supplementary Fig. 6**).

#### Exploratory Post-Hoc Contextualization of Longitudinal Integration Changes with HCs

Mean integration values were extracted from overlapping voxels that emerged across baseline and longitudinal analyses (i.e., the right anterior insula). Individuals with the greatest improvement in SOMS:CD scores showed decreases in integration from baseline to follow-up, whereas those with the greatest worsening showed modest increases, with integration values qualitatively falling predominantly within the upper and lower bounds of the HCs range (**Figure 7**). Mann-Whitney U tests revealed no significant differences in mean integration between the *entire* FND cohort and HCs at baseline (*U*=748*, p*=0.59) and follow-up (*U*=637*, p*=0.50). However, the top 20% of individuals with the greatest improvement showed significantly higher baseline integration relative to HCs (*U*=30, *p*=0.005). This difference was no longer evident at follow-up (*U*=79, *p*=0.16). In contrast, the bottom 20% of individuals with the greatest symptom worsening did not differ significantly from HCs at either baseline (*U*=88, *p*=0.24) or follow-up (*U*=103, *p*=0.45).

**Figure 7.**
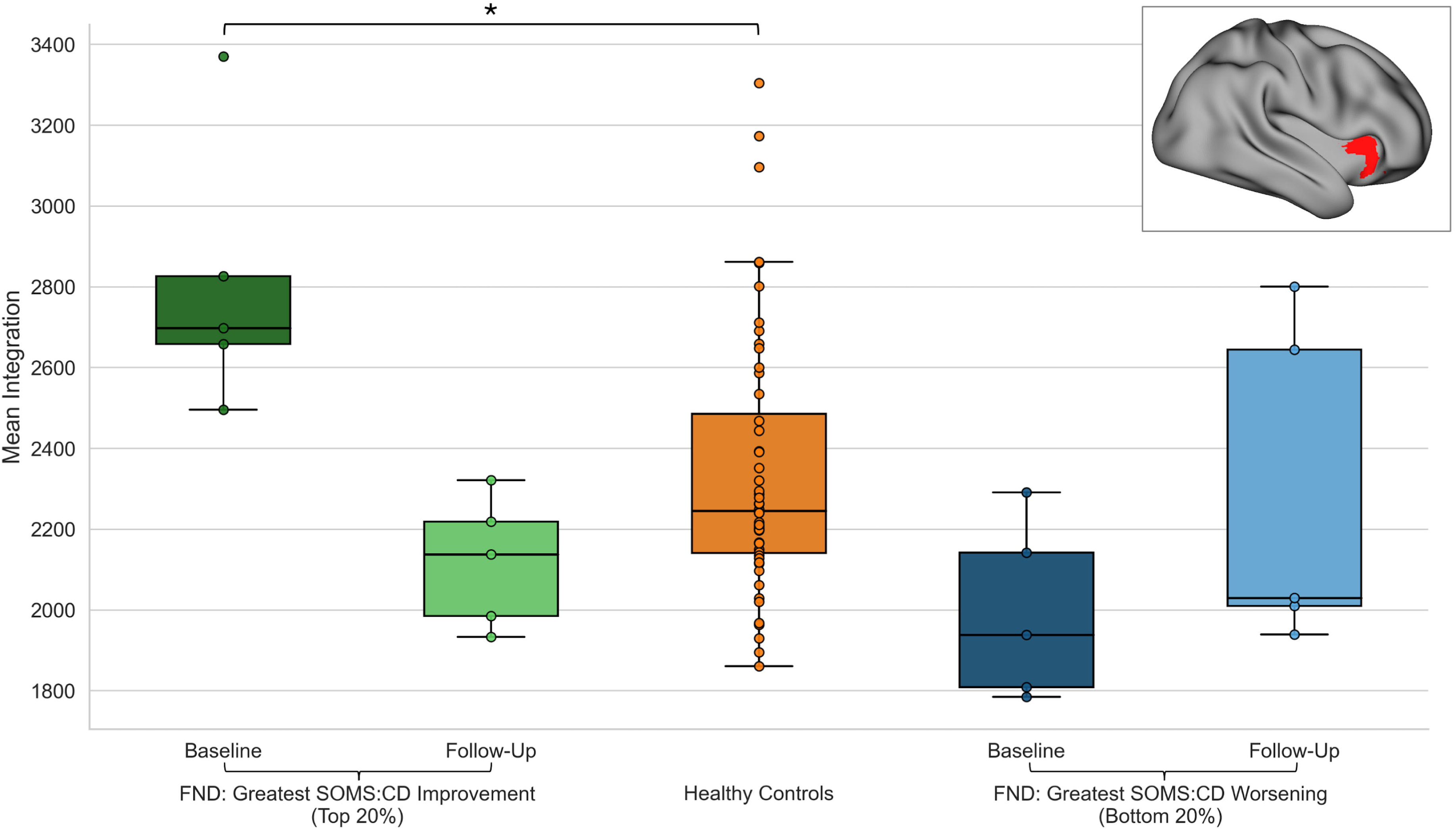
Longitudinal changes in integration-level network connectivity associated with functional neurological disorder (FND) symptom change (Δ SOMS:CD scores) in comparison to healthy controls. Boxplots depict mean integration values for overlapping voxels (shown in red; *top right* overlay image) identified across baseline predictors and longitudinal change analyses. Plots are shown for the top 20% of FND participants with the greatest symptom improvement (n=5; baseline: dark green, follow-up: light green), the bottom 20% of participants with the greatest symptom worsening (n=5; baseline: dark blue, follow-up: light blue), and healthy control participants (n=50; orange). Participants with the greatest symptom improvement exhibited decreases in mean integration values, whereas those with the greatest symptom worsening exhibited increases, with values predominantly falling within the healthy control range. Mean baseline integration values for the top five most improved participants were significantly higher than those of healthy controls (asterisk denotes *p*<0.05, Mann-Whitney U test).

#### Segregation

No regions showed an association between segregation changes and changes in SOMS:CD or PHQ-15 scores from baseline to follow-up.

## DISCUSSION

In this study, there were no group-level changes in psychometrically-measured core FND or non-core physical symptoms between baseline and follow-up; however, clinical trajectories varied considerably. Analyses using rsFC data and longitudinal psychometric scores revealed that heightened baseline connectivity in SAN, DMN, FPN, and DAN regions predicted greater symptom improvement at 6-months. Interestingly, longitudinal fMRI analyses showed that symptom improvement was associated with relative reductions in rsFC over time in many of these same networks. Overlap between predictors and mechanisms was observed for integrated (*between-network*) connectivity of the right anterior insula. Although mean integration values for the entire FND cohort in this region fell within HCs ranges at both timepoints, stratified analyses showed that participants who improved the most displayed higher baseline integration relative to HCs, which normalized over time. The divergence between null group-level findings and individual differences in network organization underscores the importance of examining within-group variability in this population. Given that most of the cohort had FND-motor, it is noteworthy that findings remained significant adjusting for FND-seizure phenotype. However, several results attenuated when controlling for baseline affective symptoms and/or trauma burden – underscoring the biological and therapeutic relevance of these factors.^40^ Together, findings demonstrate the novel observations that hyperconnectivity at baseline, and its subsequent reduction over time, are linked to more favorable clinical trajectories in FND.

When examining baseline brain organization associations with change in SOMS:CD scores, individuals with relatively increased rsFC across SAN, FPN, and DMN regions had greater longitudinal improvement in core FND symptoms. Integration analyses identified the right anterior insula and temporal pole as regions where higher between-network connectivity at baseline was related to greater improvement. The anterior insula, a core node of the SAN, has been implicated in FND pathophysiology (i.e., markers of symptom severity^17^, impaired interoceptive processing^42^), including prior research linking improvement to increased left centromedial amygdala-right anterior insula connectivity.^17^ *Post-hoc* seed-to-voxel analyses indicated that the baseline hyperconnectivity of the SAN insula seed was primarily driven by connections to the SMN, consistent with prior reports of heightened SAN-SMN coupling in FND.^21,43^ The insula cluster also overlapped with the FPN, which exhibited hyperconnectivity to both the DMN and SAN. Increased connectivity in other FPN regions (right middle frontal and precentral gyri) also predicted a favorable change in SOMS:CD scores across both weighted-degree and segregation analyses; the overlap across metrics suggests that the global centrality effects in these regions were driven by a greater number of within-FPN connections. This aligns with our prior research demonstrating increased FPN segregation in FND compared to psychiatric controls, albeit in a different subregion of the network.^21^ Overall, these results indicate that heightened connectivity at baseline – for between-network SAN, DMN, and FPN connections and within-FPN connections – is predictive of core FND symptom improvement.

Longitudinal mechanistic analyses [using fMRI and psychometric data collected across two timepoints] revealed that core FND symptom improvement was accompanied by connectivity decreases over time, consistent with the hypothesis of network reorganization accompanying symptom change. This association was particularly notable in the right anterior insula; *post-hoc* analyses indicated that reduced right insula integration reflected decreases in SAN-SMN, SAN-DMN, and FPN-DAN connectivity. When contextualizing these findings against HCs, *post-hoc* comparisons showed that right insular integration values in the *full* FND cohort did not differ from HCs at either timepoint, with values largely falling within the healthy range. In stratified exploratory *post-hoc* analyses, however, the top 20% of individuals with the greatest symptom improvement demonstrated higher baseline integration relative to HCs. This difference was no longer evident at follow-up, suggesting a shift toward a normative range. Overall, the convergence of right anterior insula findings across baseline and longitudinal analyses underscores its potential as both a prognostic marker and as a mechanistic hub whose connectivity co-varies alongside symptom improvement.

Integration analyses also identified that longitudinal connectivity decreases in the angular gyrus/TPJ and superior frontal gyrus, regions of the DMN, were linked to SOMS:CD improvement; these effects mostly reflected reduced DMN-LIM and DMN-FPN coupling. The TPJ finding is notable given its implication in FND-related action-authorship perceptions and sensorimotor integration.^19,44^ Collectively, these results indicate that clinical improvement in core FND symptoms is accompanied by targeted reductions in SAN, FPN, and DMN connectivity. These findings may help guide emerging neuromodulation approaches for FND (e.g., transcranial magnetic stimulation), by identifying cortical regions that can serve as potential targets to modulate large-scale network interactions.

For non-core physical symptoms measured by the PHQ-15, baseline analyses showed that higher segregation in the left post-central and supramarginal gyri - both within the DAN - was linked to greater improvement, whereas lower segregation in these regions was associated with worsening. Mechanistically, improvement in PHQ-15 scores corresponded to increased integration of the VIS network from baseline to follow-up, with *post-hoc* analyses showing strengthened connectivity to the FPN, DMN, and LIM networks. These results suggest that improvement in non-core physical symptoms may be supported by network patterns that are partly distinct from those underlying core FND symptoms, with an emphasis on sensory and attentional systems.^45,46^

Functional alterations in FND are increasingly being interpreted through the theoretical lens of predictive processing, which proposes that the brain draws on past experiences to actively construct predictions about incoming sensory input, rather than simply reacting to it.^47,48^ This framework has three key components, hypothesized to involve the networks showing alterations in the present study: *prediction signals* are thought to be constructed within the DMN; *prediction error signals* reflecting differences between predicted and incoming sensory inputs are thought to be precision-weighted by the SAN; and *precision signals* modulating predictions are thought to be set by the FPN.^49^ Here, we speculate that heightened baseline connectivity between the SAN and SMN may reflect a potential biasing of the system towards being overly influenced by sensorimotor signals. A reduction in SAN-SMN connectivity over time, in association with symptom improvement, may then reflect normalization of more typical interactions between these networks. Similarly, the observed increased baseline integration of the FPN with the SAN and DMN, together with increased FPN segregation, may reflect altered precision weighting that maintains maladaptive predictions. Heightened DMN connectivity that normalized with improvement may indicate a reduced persistence of maladaptive predictions. Overall, the present findings link clinical trajectories in FND to altered interactions amongst large-scale networks implicated in generating and updating predictions.

These findings and interpretations should be considered in the context of limitations. First, the naturalistic design increases ecological validity but introduces heterogeneity in treatment exposures; participants engaged in varying types, frequencies, and durations of interventions, both within and outside our healthcare system, and so we could not systematically capture all treatment details. Additionally, the observational design precludes direct causal inferences. However, as the goal of this study was to examine neural features associated with individual differences in symptom change, rather than to evaluate a specific intervention, variability in treatment is inherent to the design. Prior work from our group identified positive associations between the number of treatment sessions and FND clinical outcomes^36,37^, underscoring the importance of systematically capturing treatment details in future larger-scale research. Symptom trajectories were assessed via self-report, which is the favored approach for evaluating symptom change in FND based on international recommendations^50^; however, the inclusion of clinician-rated or objective measures of symptom change will be an important future addition. The absence of a control group of FND participants who did not receive treatment also limits our ability to estimate the natural course of symptom change. Further, several findings attenuated when accounting for baseline affective symptoms or trauma burden, indicating that they are likely components of the same complex system rather than independent factors, and should therefore be modeled as such in future research. Additionally, replication in larger FND cohorts with longer resting-state acquisitions, more controlled treatment protocols, and multimodal physiological assessments will be critical for refining and validating these network-level observations. Overall, more research is needed to fully determine if the study findings have direct mechanistic relevance or if they potentially reflect less specific features such as compensatory processes.

In conclusion, this study identified rsFC features linked to symptom change in FND. Greater baseline hyperconnectivity -- particularly involving the integrated connectivity of the right anterior insula -- predicted improvement, while recovery was characterized by reductions in these same connections. These findings highlight the potential of large-scale network interactions to serve as prognostic markers and elucidate mechanistic insights that set the stage for the development of novel, biologically-informed interventions.

## ACKNOWLEDGMENTS

We thank all the research participants, including those with FND, for their participation. This project was supported by NIMH R01MH125802.

## AUTHOR CONTRIBUTIONS

CW, CB, and DLP contributed to the conception and design of the study; CW, CB, AJG, SAF, JM, JR, JM, EG, DM, JF, CA, CDS, ID, and DLP contributed to the acquisition and analysis of data; CW, CB, and DLP contributed to drafting the text or preparing the figures.

## DATA AVAILABILITY

For qualified researchers, analysis code and de-identified data pertaining to study results can be made available following local IRB approval upon reasonable request.

## REFERENCES

1. Hallett M, Aybek S, Dworetzky BA, McWhirter L, Staab JP, Stone J. Functional neurological disorder: new subtypes and shared mechanisms. Lancet Neurol. 2022;21:537–550.

2. Jones B, Reuber M, Norman P. Correlates of health-related quality of life in adults with psychogenic nonepileptic seizures: A systematic review. Epilepsia. 2016;57:171–181.

3. Stephen CD, Fung V, Perez DL, Espay AJ. Comparison of Inpatient and Emergency Department Costs to Research Funding for Functional Neurologic Disorder: An Economic Analysis. Neurology. 2025;104:e213445.

4. Matin N, Young SS, Williams B, et al. Neuropsychiatric Associations With Gender, Illness Duration, Work Disability, and Motor Subtype in a U.S. Functional Neurological Disorders Clinic Population. J Neuropsychiatry Clin Neurosci. 2017;29:375–382.

5. Tinazzi M, Morgante F, Marcuzzo E, et al. Clinical Correlates of Functional Motor Disorders: An Italian Multicenter Study. Mov Disord Clin Pract. 2020;7:920–929.

6. McKenzie PS, Oto M, Graham CD, Duncan R. Do patients whose psychogenic non-epileptic seizures resolve, “replace” them with other medically unexplained symptoms? Medically unexplained symptoms arising after a diagnosis of psychogenic non-epileptic seizures. J Neurol Neurosurg Psychiatry. 2011;82:967–969.

7. Perez DL, Finkelstein S, Adams C, Saxena A. Toward a Precision Medicine Approach to the Outpatient Assessment and Treatment of Functional Neurological Disorder. Neurol Clin. 2023;41:681–693.

8. Sharpe M, Walker J, Williams C, et al. Guided self-help for functional (psychogenic) symptoms: a randomized controlled efficacy trial. Neurology. 2011;77:564–572.

9. Nielsen G, Stone J, Lee TC, et al. Specialist physiotherapy for functional motor disorder in England and Scotland (Physio4FMD): a pragmatic, multicentre, phase 3 randomised controlled trial. The Lancet Neurology. 2024;23:675–686.

10. Goldstein LH, Robinson EJ, Mellers JDC, et al. Cognitive behavioural therapy for adults with dissociative seizures (CODES): a pragmatic, multicentre, randomised controlled trial. Lancet Psychiatry. 2020;7:491–505.

11. Gelauff JM, Carson A, Ludwig L, Tijssen MAJ, Stone J. The prognosis of functional limb weakness: a 14-year case-control study. Brain. 2019;142:2137–2148.

12. Nielsen G, Lee TC, Marston L, et al. Which factors predict outcome from specialist physiotherapy for functional motor disorder? Prognostic modelling of the Physio4FMD intervention. Journal of Psychosomatic Research. 2025;190:112056.

13. Goldstein LH, Robinson EJ, Chalder T, et al. Moderators of cognitive behavioural therapy treatment effects and predictors of outcome in the CODES randomised controlled trial for adults with dissociative seizures. Journal of Psychosomatic Research. 2022;158:110921.

14. Sekine ER, Kanaan RA, McMillan J, Oxford S, Iles RA. Biopsychosocial prognostic indicators in Functional Neurological Disorder: A systematic review. J Psychosom Res. 2025;195:112201.

15. Perez DL, Nicholson TR, Asadi-Pooya AA, et al. Neuroimaging in Functional Neurological Disorder: State of the Field and Research Agenda. Neuroimage Clin. 2021;30:102623.

16. Baek K, Doñamayor N, Morris LS, et al. Impaired awareness of motor intention in functional neurological disorder: implications for voluntary and functional movement. Psychological Medicine. 2017;47:1624–1636.

17. Diez I, Ortiz-Terán L, Williams B, et al. Corticolimbic fast-tracking: enhanced multimodal integration in functional neurological disorder. J Neurol Neurosurg Psychiatry. 2019;90:929–938.

18. Li R, Li Y, An D, Gong Q, Zhou D, Chen H. Altered regional activity and inter-regional functional connectivity in psychogenic non-epileptic seizures. Sci Rep. 2015;5:11635.

19. Maurer CW, LaFaver K, Ameli R, Epstein SA, Hallett M, Horovitz SG. Impaired self-agency in functional movement disorders. Neurology. 2016;87:564–570.

20. van der Kruijs SJM, Bodde NMG, Vaessen MJ, et al. Functional connectivity of dissociation in patients with psychogenic non-epileptic seizures. J Neurol Neurosurg Psychiatry. 2012;83:239–247.

21. Westlin C, Guthrie AJ, Bleier C, et al. Delineating network integration and segregation in the pathophysiology of functional neurological disorder. Brain Communications. 2025;7:fcaf195.

22. Conejero I, Collombier L, Lopez-Castroman J, et al. Association between brain metabolism and clinical course of motor functional neurological disorders. Brain. 2022;145:3264–3273.

23. Espay AJ, Ries S, Maloney T, et al. Clinical and neural responses to cognitive behavioral therapy for functional tremor. Neurology. 2019;93:e1787–e1798.

24. Faul L, Knight LK, Espay AJ, Depue BE, LaFaver K. Neural activity in functional movement disorders after inpatient rehabilitation. Psychiatry Research: Neuroimaging. 2020;303:111125.

25. Schneider A, Weber S, Wyss A, Loukas S, Aybek S. BOLD signal variability as potential new biomarker of functional neurological disorders. NeuroImage: Clinical. 2024;43:103625.

26. Szaflarski JP, LaFrance WC, Nenert R, et al. Neurobehavioral therapy in functional seizures: Investigation of mechanism of action with resting-state functional magnetic resonance imaging. Epilepsia. 2025; 66(8):2881–2893.

27. LaFrance WC, Baker GA, Duncan R, Goldstein LH, Reuber M. Minimum requirements for the diagnosis of psychogenic nonepileptic seizures: a staged approach: a report from the International League Against Epilepsy Nonepileptic Seizures Task Force. Epilepsia. 2013;54:2005–2018.

28. Aybek S, Perez DL. Diagnosis and management of functional neurological disorder. BMJ. 2022;376:o64.

29. Westlin C, Guthrie AJ, Paredes-Echeverri S, et al. Machine learning classification of functional neurological disorder using structural brain MRI features. J Neurol Neurosurg Psychiatry. 2025;96:249–257.

30. Westlin C, Guthrie A, Bleier C, et al. Functional connectivity gradients reveal altered hierarchical cortical organization in functional neurological disorder. Biological Psychiatry: Cognitive Neuroscience & Neuroimaging. 2025; Oct 28:S2451-9022(25)00325-8.

31. Myers L, Sarudiansky M, Korman G, Baslet G. Using evidence-based psychotherapy to tailor treatment for patients with functional neurological disorders. Epilepsy Behav Rep. 2021;16:100478.

32. Baker J, Barnett C, Cavalli L, et al. Management of functional communication, swallowing, cough and related disorders: consensus recommendations for speech and language therapy. J Neurol Neurosurg Psychiatry. 2021;92:1112–1125.

33. Nicholson C, Edwards MJ, Carson AJ, et al. Occupational therapy consensus recommendations for functional neurological disorder. J Neurol Neurosurg Psychiatry. 2020;91:1037–1045.

34. Nielsen G, Stone J, Matthews A, et al. Physiotherapy for functional motor disorders: a consensus recommendation. J Neurol Neurosurg Psychiatry. 2015;86:1113–1119.

35. LaFrance WC Jr, Baird GL, Barry JJ, et al. Multicenter Pilot Treatment Trial for Psychogenic Nonepileptic Seizures: A Randomized Clinical Trial. JAMA Psychiatry. 2014;71:997–1005.

36. Bleier C, Godena E, Millstein D, et al. Predictors of Skills-Based Psychotherapy Outcomes for Functional Neurological Disorder: A Retrospective Cohort Study. J Neuropsychiatry Clin Neurosci. Epub 2025.

37. Maggio JB, Ospina JP, Callahan J, Hunt AL, Stephen CD, Perez DL. Outpatient Physical Therapy for Functional Neurological Disorder: A Preliminary Feasibility and Naturalistic Outcome Study in a U.S. Cohort. J Neuropsychiatry Clin Neurosci. 2020;32:85–89.

38. Rief W, Hiller W. A new approach to the assessment of the treatment effects of somatoform disorders. Psychosomatics. 2003;44:492–498.

39. Kroenke K, Spitzer RL, Williams JBW. The PHQ-15: validity of a new measure for evaluating the severity of somatic symptoms. Psychosom Med. 2002;64:258–266.

40. Diez I, Larson AG, Nakhate V, et al. Early-life trauma endophenotypes and brain circuit–gene expression relationships in functional neurological (conversion) disorder. Mol Psychiatry. 2021;26:3817–3828.

41. Yeo BTT, Krienen FM, Sepulcre J, et al. The organization of the human cerebral cortex estimated by intrinsic functional connectivity. Journal of Neurophysiology. 2011;106:1125.

42. Sojka P, Serranová T, Khalsa SS, Perez DL, Diez I. Altered Neural Processing of Interoception in Patients With Functional Neurological Disorder: A Task-Based fMRI Study. J Neuropsychiatry Clin Neurosci. 2025; 37(2):149–159.

43. Voon V, Brezing C, Gallea C, et al. Emotional stimuli and motor conversion disorder. Brain. 2010;133:1526–1536.

44. Voon V, Gallea C, Hattori N, Bruno M, Ekanayake V, Hallett M. The involuntary nature of conversion disorder. Neurology. 2010;74:223–228.

45. Věchetová G, Nikolai T, Slovák M, et al. Attention impairment in motor functional neurological disorders: a neuropsychological study. J Neurol. 2022;269:5981–5990.

46. Huys A-CML, Haggard P, Bhatia KP, Edwards MJ. Misdirected attentional focus in functional tremor. Brain. 2021;144:3436–3450.

47. Edwards MJ, Adams RA, Brown H, Pareés I, Friston KJ. A Bayesian account of ‘hysteria.’ Brain. 2012;135:3495–3512.

48. Jungilligens J, Paredes-Echeverri S, Popkirov S, Barrett LF, Perez DL. A new science of emotion: implications for functional neurological disorder. Brain. 2022;145:2648–2663.

49. Barrett LF. The theory of constructed emotion: an active inference account of interoception and categorization. Soc Cogn Affect Neurosci. 2017;12:1–23.

50. Pick S, Anderson DG, Asadi-Pooya AA, et al. Outcome measurement in functional neurological disorder: a systematic review and recommendations. J Neurol Neurosurg Psychiatry. 2020;91:638–649.

