## Supplementary Materials for "Functional Connectivity Predictors and Mechanisms of Symptom Change in Functional Neurological Disorder"

**MRI Acquisition and Preprocessing**

Participants were scanned on a Siemens Tim Trio 3T MRI scanner using a 12-channel phased-array head coil. A high-resolution T1-weighted magnetization-prepared rapid gradient echo (MP-RAGE) scan was acquired for each subject with the following parameters: 1mm isotropic voxels; 160 sagittal slices; acquisition matrix size=256x256; repetition time=2300ms; echo time=2.98ms; field of view=256mm. Resting-state blood-oxygen-level-dependent functional scans were acquired using T2*-weighted echo-planar imaging sequences with the following parameters: TR=3000ms; TE=30ms; flip angle 85; 216mm FOV; 3mm isotropic voxels; sequence length=6 minutes, 12 seconds (124 time points/scan). All participants had two functional acquisitions. Participants were instructed to remain as still as possible with their eyes open, bi-temporal foam pads restricted head motion, and earplugs were used to attenuate scanner noise.

Preprocessing for anatomical and functional MRI data was done using FMRIB Software Library v5.0.7 (FSL, Oxford, UK) and MATLAB 2023a (MathWorks, Natick, MA) with in-house preprocessing pipelines that have been described previously (Diez et al., 2021; Westlin et al., 2025). Preprocessing of T1-weighted anatomical images included: reorientation to right-posterior-inferior (RPI); alignment to the anterior and posterior commissures; skull stripping; segmentation of grey matter, white matter, and cerebrospinal fluid; and computation of non-linear transformation between individual skull-stripped T1 images and a 3mm resolution MNI152 template. fMRI preprocessing steps included: discarding the first four volumes; slice timing correction; reorientation to RPI; realignment of functional volumes via a 6-parameter rigid body transformation; computation of the transformation between individual skull-stripped T1 images and mean functional images; intensity normalization; and regression of nuisance signals, including 12 motion-related covariates (rigid motion parameters and their derivatives), linear and quadratic terms, and five components each from the lateral ventricles and white matter. Further steps included transformation to MNI space, spatial smoothing using a 6mm full-width at half-maximum Gaussian kernel, and temporal band-pass filtering (0.01-0.08Hz). Head motion was quantified using realignment parameters, including three translation and three rotation estimates, and volumes with excessive head motion (framewise displacement >0.5mm) were removed from the data. Analyses were performed on 120 time points per subject; resting-state acquisition runs were concatenated, and the first 120 time points were retained to standardize the time series length across participants.

**Laboratory Designed Follow-up Questions**

1). In the past 6 months, have you participated in physical therapy?

Yes	No

1.a). If yes, how many sessions did you participate in over the past 6 months?

0-5

6-10

11-15

16-20

>20

2). In the past 6 months, have you participated in occupational therapy?

Yes	No

2.a). If yes, how many sessions did you participate in over the past 6 months?

0-5

6-10

11-15

16-20

>20

3). In the past 6 months, have you participated in speech and language therapy?

Yes	No

3.a). If yes, how many sessions did you participate in over the past 6 months?

0-5

6-10

11-15

16-20

>20

4). In the past 6 months, have you participated in individual psychotherapy?

Yes	No

4.a). If yes, how many sessions did you participate in over the past 6 months?

0-5

6-10

11-15

16-20

>20

4.b). If yes, are you participating in cognitive behavioral therapy?

Yes	No

5). In the past 6 months, have you participated in group psychotherapy?

Yes	No

5.a). If yes, how many sessions did you participate in over the past 6 months?

0-5

6-10

11-15

16-20

>20

6). In the last 6 months, did you use in any capacity the workbook "Overcoming Functional Neurological Symptoms: A Five Area Approach"?

Yes	No

7). In the last 6 months, did you use in any capacity the workbook "Taking Control of Your Seizures: Workbook (Treatments That Work)"?

Yes	No

8). Select below the option that best describes your functional neurological disorder treatment.

- All treatment within the Mass General Brigham system

- Some community and some Mass General Brigham system

- No treatment received within the Mass General Brigham system

9). Are you currently (Check all that apply):

- Unemployed

- Student

- Homemaker

- Working part time

- Working full time

- Retired

- On disability

- Applying for disability

- Other

10). Are you currently (Check all that apply):

- Single

- In a relationship

- Engaged

- Married

- Divorced

- Widowed

- Other

**SUPPLEMENTARY TABLES**

**Supplementary Table 1. Demographic characteristics of participants with functional neurological disorder (FND).**

| **FND Subject** | **FND**  **Phenotype** | **Phenotype Descriptions** | **Current (Baseline) SCID-DSM-5 Diagnoses** | **Past SCID-DSM-5 Diagnoses** | **Baseline**  **Psychotropic Medications** | **Follow-up**  **Psychotropic Medications** |
| --- | --- | --- | --- | --- | --- | --- |
| 1 | FND-Motor | clinically-established functional tremor & functional gait | MDE, SAD | MDE, PD+AG | FLX, CLP | DLX, GBP, CLP |
| 2 | FND-Motor | clinically-established functional gait & functional speech | PTSD, Eating Disorder | MDE | QTP, ECP, CLP, HDZ, PZN | VNF, QTP, HDZ, PZN, CLP |
| 3 | FND-Seizure | documented functional seizures | DEP NOS | MDE, AG | NRT, LTG | LTG, PZN, NRT, HDZ |
| 4 | FND-Motor** | clinically-established functional tremor | GAD, IAD | MDE, PTSD | DLX, LRZ | LRZ, DLX |
| 5* | FND-Motor | clinically-established functional jerky movements | GAD | Eating Disorder, AUD, SUD | ECP, PGB, AMT, APM / DXAM | ECP |
| 6 | FND-Motor | clinically-established functional tremor | ANX NOS | - | GBP | GBP |
| 7* | FND-Seizure | documented functional seizures | - | DEP NOS, SAD, ANX NOS | LRZ | LRZ |
| 8 | FND-Seizure | probable functional seizures | ANX NOS | PTSD, GAD, MDE | SERT, TZD, PZN | SERT, TZD |
| 9 | FND-Motor | clinically-established functional jerky movements | GAD | DEP NOS | SERT | SERT |
| 10 | FND-Motor | clinically-established functional limb weakness (left arm) & functional speech | GAD, SSD | MDE, PTSD | CBD | - |
| 11 | FND-Seizure | probable functional seizures | - | ANX NOS | - | - |
| 12 | FND-Seizure, FND-Motor | clinically-established functional tremor, probable functional seizures | - | MDE | AMT | AMT |
| 13 | FND-Motor** | clinically-established functional tremor & functional speech | PTSD, GAD, PD+AG, MDE | - | DLX, BUP, LRZ | DLX, BUP, LRZ |
| 14 | FND-Motor | clinically-established functional gait & functional speech | AG, ADHD | - | APZ, CBD | APZ |
| 15 | FND-Seizure, FND-Motor | documented functional seizures; clinically-established functional gait & functional speech | ANX NOS | PTSD, PD+AG | PGB, CBD | PGB |
| 16 | FND-Motor | clinically-established functional gait, functional tremor, functional limb weakness (right arm/leg), & functional speech | GAD, PD+AG | PTSD, MDE | TPM | TPM |
| 17 | FND-Motor** | clinically-established functional gait | ANX NOS, SSD | GAD, MDE | SERT, LRZ, GBP | SERT, LRZ, GBP |
| 18 | FND-Motor | clinically-established functional tremor | - | PTSD, BPAD-II (with MDE) | LTM, QTP, LRZ | LTM, QTP, LRZ |
| 19 | FND-Motor | clinically-established functional limb weakness (legs) & functional speech | - | DEP NOS, Specific Phobia | - | - |
| 20 | FND-Motor | clinically-established functional tremor, functional dystonia & functional gait | PD+AG, SSD | GAD, DEP NOS | - | ATX |
| 21 | FND-Motor | clinically-established functional facial spasms/tics & functional speech | GAD, PD+AG, ADHD, PTSD | MDE | CLN, LDA, APR, SERT, LRZ | CLN, LDA, SERT |
| 22 | FND-Motor | clinically-established functional gait & functional speech | - | DEP NOS, ANX NOS | GBP | GBP |
| 23 | FND-Motor | clinically-established functional limb weakness (left leg) | ANX NOS | MDE, ANX NOS | GBP, HDZ, ECP | ECP, DLX, HDZ |
| 24 | FND-Seizure, FND-Motor | probable functional seizures; clinically-established functional limb weakness (bilateral leg), functional tremor, & functional gait | SSD, AG, DYS | MDE, PD-AG, GAD, PTSD, Eating Disorder | AMT, DLX | AMT, DLX |
| 25 | FND-Motor | clinically-established functional dystonia & functional speech | GAD | PTSD | - | - |
| 26 | FND-Motor | clinically-established functional limb weakness (left arm & leg), functional tremor, & functional jerks | - | PTSD | - | - |
| 27 | FND-Motor | clinically-established functional tremor | PTSD, MDE, Anxiety NOS | - | MIR, CLP | MIR, CLP |
| 28 | FND-Seizure, FND-Motor | documented functional seizures; clinically-established functional jerks/tics, & functional speech | PD+AG, DYS, GAD, SSD | OCD | DLX, GBP, LRZ | AMA, VNF, HDZ |
| 29* | FND-Seizure | documented functional seizures | AG, SAD, OCD, SSD | - | HDZ, DZP | HDZ, DZP, SERT |
| 30* | FND-Motor | clinically-established functional tremor, & functional gait | Anxiety NOS | - | HDZ | - |
| 31 | FND-Seizure, FND-Motor | documented functional seizures; clinically-established functional gait, functional limb weakness (left arm and leg), & functional speech | PTSD, MDE, PD+AG, GAD, SSD | MDE, Eating Disorder, AUD, SUD | BUP, GBP, TZD, HDZ | BUP, GBP, HDZ, TZD |
| 32 | FND-Motor | clinically-established functional limb weakness (left arm and leg) & functional gait | - | MDE, SAD, PTSD, Eating Disorder | CTP | CTP |

*Indicates subjects who did not have a scan at the 6-month follow-up session. **Indicates subject also had concurrent functional somatosensory loss (e.g., non-dermatomal somatosensory deficits). FND-Motor, Functional Motor Disorder; FND-Seizure, Functional Seizures; SCID-DSM-5, Structured Clinical Interview for DSM 5^th^ Edition [note subject 1 received a SCID-I for DSM-IV]; ADHD, Attention-Deficit/Hyperactivity Disorder; AG, Agoraphobia; ANX, Anxiety; AUD, Alcohol Use Disorder; BPAD, Bipolar Affective Disorder; DEP, Depression; DYS, Dysthymia; GAD, Generalized Anxiety Disorder; IAD, Illness Anxiety Disorder; MDE, Major Depressive Episode; NOS, not otherwise specified; OCD, Obsessive Compulsive Disorder; PD+AG, Panic Disorder with Agoraphobia; PD-AG, Panic Disorder without Agoraphobia; PTSD, Post-Traumatic Stress Disorder; SAD, Social Anxiety Disorder; SSD, Somatic Symptom Disorder; SUD, Substance Use Disorder; AMA, Amantadine; AMT, Amitriptyline; APM, Amphetamine; APR, Aripiprazole; APZ, Alprazolam; ATX, Atomoxetine; BUP, Bupropion; CBD, Cannabidiol; CLN, Clonidine; CLP, Clonazepam; CTP, Citalopram; DLX, Duloxetine; DXAM, Dextroamphetamine; DZP, Diazepam; ECP, Escitalopram; FLX, Fluoxetine; GBP, Gabapentin; HDZ, Hydroxyzine; LDA, Lisdexamfetamine; LTM, Lithium; LTG, Lamotrigine; LRZ, Lorazepam; MIR, Mirtazapine; NRT, Nortriptyline; PGB, Pregabalin; PZN, Prazosin; QTP, Quetiapine; SERT, Sertraline; TPM, Topiramate; TZD, Trazodone; VNF, Venlafaxine.

**Supplementary Table 2. Baseline characteristics of participants who completed vs. did not complete 6-month follow-up sessions.**

|  | **Participants who completed follow-up (n=32)** Mean ± SD or n (%) | **Participants lost to follow-up (n=20)**  Mean ± SD or n (%) | **Test statistic** | **Uncorrected**  **p-value** | **FDR corrected p-value** |
| --- | --- | --- | --- | --- | --- |
| **Age at consent (yrs.)** | 42.4 ± 13.3; range: 22.4-72.1; median: 44.0 | 37.1 ± 13.8; range: 19.6-65.8; median: 33.9 | 1.24^a^ | 0.17 | 0.44 |
| **Biological sex** | 28F 4M | 17F 3M | 0.07^b^ | 0.80 | 0.80 |
| **Illness duration (yrs.)** | 3.7 ± 4.2; range: 0.1-16.1; median: 2.1 | 2.3 ± 1.4; range: 0.5-5.4; median: 1.8 | 325.0^c^ | 0.46 | 0.66 |
| **Race (white)** | 28 (87.5%) | 17 (85.0%) | 0.07^b^ | 0.79 | 0.80 |
| **Married** | 15 (46.9%) | 8 (40.0%) | 0.24^b^ | 0.63 | 0.74 |
| **College graduate** | 22 (68.8%) | 10 (50.0%) | 1.00^b^ | 0.32 | 0.57 |
| **Employed (or student full time)** | 12 (37.5%) | 5 (25.0%) | 0.87^b^ | 0.35 | 0.57 |
| **On medical disability** | 10 (31.3%) | 8 (40.0%) | 0.32^b^ | 0.32 | 0.57 |
| **Functional motor disorder** | 27 (84.4%) | 11 (55.0%) | 3.90^b^ | 0.05 | 0.21 |
| **Functional seizures** | 10 (31.3%) | 8 (40.0%) | 0.42^b^ | 0.52 | 0.67 |
| **Taking SSRI/SNRI at baseline** | 13 (40.6%) | 12 (60.0%) | 2.93^b^ | 0.09 | 0.28 |
| **Baseline SOMS:CD** | 6.6 ± 6.3, range: 0-21; median: 5 | 12.2 ± 8.7, range: 0-28; median: 11 | 234.5^c^ | 0.03 | 0.21 |
| **Baseline STAI-state** | 38.3 ± 11.6, range: 20-68; median: 35 | 42.6 ± 8.9, range: 31-65; median: 39 | 237.5^c^ | 0.05 | 0.21 |

^a^ Test statistic corresponds to t-test. ^b^ Test statistic corresponds to chi-square test. ^c^ Test statistic corresponds to Mann-Whitney U test. SSRI, Selective Serotonin Reuptake Inhibitor; SNRI, Serotonin and Norepinephrine Reuptake Inhibitor, SOMS:CD Screening for Somatoform Symptoms-7 Subscale for Conversion Disorder; STAI-state, Spielberger State-Trait Anxiety Inventory-State subscale.

**Supplementary Table 3. Demographic and psychometric characteristics of healthy control (HC) cohort.**

|  | **HCs (n=59)**  Mean ± SD or n |
| --- | --- |
| **Age at consent (yrs.)** | 36.6 ± 10.9 |
| **Biological sex** | F:48 M:11 |
| **SOMS:CD** | 0 ± 0 |
| **PHQ-15** | 2.6 ± 2.2 |
| **BDI-II** | 1.5 ± 2.5 |
| **STAI-Total** | 53.9 ± 9.3 |
| **PCL-5** | 2.7 ± 3.6 |
| **CTQ-abuse** | 17.9 ± 3.4 |
| **CTQ-neglect** | 13.2 ± 4.3 |

SOMS:CD, Screening for Somatoform Symptoms-7 Subscale for Conversion Disorder; PHQ-15, Patient Health Questionnaire-15; BDI-II, Beck Depression Inventory-II; STAI, Spielberg State-Trait Anxiety Inventory; PCL-5, PTSD Checklist 5; CTQ, Childhood Trauma Questionnaire.

**Supplementary Table 4.** **Responses to follow-up questionnaire assessing therapy participation, treatment setting, and sociodemographic factors.**

| **Laboratory Designed Questions** | **Responses (n=32)** |
| --- | --- |
| 1. Physical therapy | Yes: 17, No: 15 |
| 1.a. Physical therapy sessions | (0-5): 4  (6-10): 3  (11-15): 1  (16-20): 2  (>20): 7 |
| 2. Occupational therapy^a^ | Yes: 16^a^, No: 16 |
| 2.a. Occupational therapy sessions^a^ | (0-5): 6  (6-10): 6  (11-15): 1  (16-20): 0  (>20): 3 |
| 3. Speech and language therapy^a^ | Yes: 10, No: 22 |
| 3.a. Speech and language therapy sessions^a^ | (0-5): 7  (6-10): 3  (11-15): 0  (16-20): 0  (>20): 0 |
| 4. Individual psychotherapy^a^ | Yes: 29, No: 3 |
| 4.a. Individual psychotherapy sessions^a^ | (0-5): 8  (6-10): 6  (11-15): 5  (16-20): 10  (>20): 0 |
| 4.b. Cognitive-behavioral therapy^a^ | Yes: 20, No: 9 |
| 5. Group psychotherapy^a^ | Yes: 5, No: 27 |
| 5.a. Group psychotherapy sessions^a^ | (0-5): 0  (6-10): 4  (11-15): 0  (16-20): 1  (>20): 0 |
| 6. Used workbook *“Overcoming Functional Neurological Symptoms: A Five Area Approach”*? | Yes: 17, No: 15 |
| 7. Used workbook *“Taking Control of Your Seizures: Workbook (Treatments That Work)”*? | Yes: 5, No: 27 |
| 8. FND treatment setting | - All treatment within the Mass General Brigham system: 11  - Some community and some Mass General Brigham system: 16  - No treatment received within the Mass General Brigham system: 5 |
| 9. Employment status (all that apply) | Unemployed 5  Student 3  Homemaker 2  Working part time 10  Working full time 14  Retired 0  On disability 7  Applying for disability 5  Other 1 |
| 10. Relationship status (all that apply) | Single 7  In a relationship 7  Engaged 1  Married 15  Divorced 4  Widowed 2  Other 0 |

^a^ Two participants initially self-reported not receiving psychotherapy in the 6 months prior to the follow-up questionnaire. However, cross-reference with available medical records revealed that they had both participated in individual psychotherapy and one of them had also participated in group psychotherapy during that period. In total, 30 of 32 individuals received some form of psychotherapy (individual and/or group) in between their baseline and follow-up assessments; 21 of 32 received a component of cognitive behavioral therapy (individual and/or group) in their psychotherapy. Similarly, two participants underreported receiving occupational therapy, and one underreported receiving speech and language therapy. Counts presented reflect the chart-verified (corrected) values.

**Supplementary Table 5. Relationship between baseline clinical characteristics and symptom change [follow-up vs. baseline] in the longitudinal functional neurological disorder cohort.**

| **Outcome**  **Variable** | **Baseline**  **Variable** | **Test**  **Statistic** | **Uncorrected**  ***p*-value** | **FDR- corrected**  ***p*-value** |
| --- | --- | --- | --- | --- |
| **SOMS:CD Change** |  |  |  |  |
|  | Age at consent | 0.008^a^ | 0.964 | 1.000 |
|  | Biological sex | 56.500^b^ | 1.000 | 1.000 |
|  | Illness duration | -0.329^a^ | 0.066 | 0.640 |
|  | Interval follow-up period | 0.220^a^ | 0.226 | 0.640 |
|  | Race (white) | 84.500^b^ | 0.106 | 0.640 |
|  | Married | 129.500^b^ | 0.954 | 1.000 |
|  | College graduate | 95.000^b^ | 0.550 | 0.946 |
|  | Employed full-time (or full-time student) | 96.000^b^ | 0.354 | 0.860 |
|  | On medical disability or worker's compensation | 107.500^b^ | 0.934 | 1.000 |
|  | Functional motor disorder | 59.000^b^ | 0.674 | 0.955 |
|  | Functional seizures | 148.000^b^ | 0.123 | 0.640 |
|  | SSRI/SNRI use | 106.000^b^ | 0.509 | 0.946 |
|  | Baseline BDI-II | -0.023^a^ | 0.902 | 1.000 |
|  | Baseline STAI-total | 0.098^a^ | 0.595 | 0.946 |
|  | Baseline PCL-5 | 0.093^a^ | 0.612 | 0.946 |
|  | Baseline CTQ-abuse | -0.237^a^ | 0.191 | 0.640 |
|  | Baseline CTQ-neglect | -0.230^a^ | 0.206 | 0.640 |
| **PHQ-15 Change** |  |  |  |  |
|  | Age at consent | 0.001^c^ | 0.994 | 0.994 |
|  | Biological sex | 0.225^d^ | 0.824 | 0.984 |
|  | Illness duration | 0.004^a^ | 0.982 | 0.994 |
|  | Interval follow-up period | 0.138^a^ | 0.451 | 0.982 |
|  | Race (white) | 0.266^d^ | 0.792 | 0.984 |
|  | Married | 0.451^d^ | 0.655 | 0.984 |
|  | College graduate | -0.168^d^ | 0.868 | 0.984 |
|  | Employed full-time (or full-time student) | 1.155^d^ | 0.257 | 0.945 |
|  | On medical disability or worker's compensation | 66.000^b^ | 0.075 | 0.945 |
|  | Functional motor disorder | 1.105^d^ | 0.278 | 0.945 |
|  | Functional seizures | 81.500^b^ | 0.252 | 0.945 |
|  | SSRI/SNRI use | 128.500^b^ | 0.862 | 0.984 |
|  | Baseline BDI-II | 0.135^a^ | 0.462 | 0.982 |
|  | Baseline STAI-total | 0.035^c^ | 0.851 | 0984 |
|  | Baseline PCL-5 | 0.152^a^ | 0.405 | 0.982 |
|  | Baseline CTQ-abuse | 0.250^a^ | 0.167 | 0.945 |
|  | Baseline CTQ-neglect | 0.101^a^ | 0.583 | 0.984 |
|  | Baseline CTQ-neglect | 106.500^b^ | 0.525 | 1.000 |
| **CGI-I Improved vs. Not Improved** |  |  |  |  |
|  | Age at consent | -0.424^d^ | 0.675 | 1.000 |
|  | Biological sex | 0.647^e^ | 1.000 | 1.000 |
|  | Illness duration | 177.000^b^ | 0.042* | 0.417 |
|  | Interval follow-up period | 137.000^b^ | 0.618 | 1.000 |
|  | Race (white) | 0.444^e^ | 0.629 | 1.000 |
|  | Married | 0.005^f^ | 0.946 | 1.000 |
|  | College graduate | 1.750^e^ | 0.699 | 1.000 |
|  | Employed full-time (or full-time student) | 0.933^e^ | 1.000 | 1.000 |
|  | On medical disability or worker's compensation | 1.038^e^ | 1.000 | 1.000 |
|  | Functional motor disorder | 2.550^e^ | 0.374 | 1.000 |
|  | Functional seizures | 0.161^e^ | 0.049* | 0.417 |
|  | SSRI/SNRI use | 0.042^f^ | 0.837 | 1.000 |
|  | Baseline BDI-II | 127.500^b^ | 0.893 | 1.000 |
|  | Baseline STAI-total | 0.929^d^ | 0.360 | 1.000 |
|  | Baseline PCL-5 | 124.500^b^ | 0.985 | 1.000 |
|  | Baseline CTQ-abuse | 101.500^b^ | 0.409 | 1.000 |
|  | Baseline CTQ-neglect | 106.500^b^ | 0.525 | 1.000 |

*indicates uncorrected p-value <0.05. ^a^Test statistic corresponds to Spearman correlation. ^b^Test statistic corresponds to Mann-Whitney U test. ^c^Test statistic corresponds to Pearson correlation. ^d^Test statistic corresponds to T-test. ^e^Test statistic corresponds to Fisher’s exact test. ^f^Test statistic corresponds to chi-squared test. CGI-I improved includes “much improved” and “improved”, while CGI-I not-improved “unchanged” and “worse,” based on patient-reported Clinical Global Impression of Improvement (CGI-I) scale scores. False discovery rate (FDR) correction was performed across all baseline variables for a given symptom outcome measure. SOMS:CD, Screening for Somatoform Symptoms-7 Subscale for Conversion Disorder; PHQ-15, Patient Health Questionnaire-15; BDI-II, Beck Depression Inventory-II; STAI, Spielberg State-Trait Anxiety Inventory; PCL-5, PTSD Checklist 5; CTQ, Childhood Trauma Questionnaire.

**SUPPLEMENTARY FIGURE CAPTIONS**

**Supplementary Fig. 1.** Upset plot of functional neurological symptoms across the functional neurological disorder (FND) cohort. Vertical bars reflect the number of individuals with each combination of co-occurring symptoms, as shown by multiple darkened circles connected by lines, or single symptom presentation indicated by one darkened circle. Horizontal bars reflect the total number of individuals who experienced each individual symptom (with or without other co-occurring symptoms).

**Supplementary Fig. 2.** Results from analyses relating functional neurological disorder (FND) symptom change scores to baseline resting-state functional connectivity metrics (*left,* weighted-degree; *middle,* integration; *right*, segregation). The top row depicts results from the primary adjustment for age, sex, SSRI/SNRI use, mean framewise displacement (FD), elapsed time between baseline and follow-up sessions, and baseline SOMS:CD score; the second row depicts results from the *post-hoc* adjustment controlling additionally for FND phenotype (FND-seizure yes/no); the third row depicts results from the *post-hoc* adjustment controlling additionally for baseline BDI-II, STAI-total, and PCL-5 scores; the bottom row depicts results from the *post-hoc* adjustment controlling additionally for CTQ-abuse and CTQ-neglect scores. Colors reflect the z-statistic for the corresponding general linear (z-statistic>1.96; p<0.05 cluster-corrected for multiple comparisons). Volumetric brain maps are visualized on the surface for display purposes only; subcortical results in volume are shown with the left hemisphere on the left side of the image. SOMS:CD, Screening for Somatoform Symptoms-7 Subscale for Conversion Disorder; SSRI/SNRI, selective serotonin/norepinephrine reuptake inhibitor; FD, framewise displacement; BDI-II, Beck Depression Inventory-II; STAI-total, State Trait Anxiety Inventory-Total; PCL-5, PTSD Checklist-5; CTQ, Childhood Trauma Questionnaire.

**Supplementary Fig. 3.** Network connectivity of baseline integration predictors associated with functional neurological disorder (FND) symptom change. Only three networks containing the most significant voxels are displayed: (**A**) salience network; (**B**) frontoparietal network; (**C**) default mode network. For each of these networks, we extracted the average time series of all significant voxels for use in seed-to-voxel connectivity analyses, to then characterize the networks they were functionally integrating across. Pie charts detail the percent of significant connections from the seed network to each of the other Yeo et al. (2011) networks. Self-connections (e.g., DMN-to-DMN) were excluded, as integration analyses focused on cross-network connectivity. Charts excluded networks with fewer than 1.0% of connections.

**Supplementary Fig. 4.** Results from analyses relating physical symptom change scores to baseline resting-state functional connectivity metrics (*left,* weighted-degree; *middle,* integration; *right*, segregation). The top row depicts results from the primary adjustment for age, sex, SSRI/SNRI use, mean framewise displacement (FD), elapsed time between baseline and follow-up sessions, and baseline PHQ-15 score; the second row depicts results from the *post-hoc* adjustment controlling additionally for FND phenotype (FND-seizure yes/no); the third row depicts results from the *post-hoc* adjustment controlling additionally for baseline BDI-II, STAI-total, and PCL-5 scores; the bottom row depicts results from the *post-hoc* adjustment controlling additionally for CTQ-abuse and CTQ-neglect scores. Colors reflect the z-statistic for the corresponding general linear (z-statistic>1.96; p<0.05 cluster-corrected for multiple comparisons). Volumetric brain maps are visualized on the surface for display purposes only. PHQ-15, Patient Health Questionnaire-15; SSRI/SNRI, selective serotonin/norepinephrine reuptake inhibitor; FD, framewise displacement; BDI-II, Beck Depression Inventory-II; STAI-total, State Trait Anxiety Inventory-Total; PCL-5, PTSD Checklist-5; CTQ, Childhood Trauma Questionnaire.

**Supplementary Fig. 5.** Results from analyses relating FND symptom change scores to changes in resting-state functional connectivity metrics (*left,* weighted-degree; *middle,* integration; *right*, segregation) between follow-up and baseline. The top row depicts results from the primary adjustment for age, sex, SSRI/SNRI use, mean framewise displacement (FD), elapsed time between baseline and follow-up sessions, and baseline SOMS:CD score; the second row depicts results from the *post-hoc* adjustment controlling additionally for FND phenotype (FND-seizure yes/no); the third row depicts results from the *post-hoc* adjustment controlling additionally for baseline BDI-II, STAI-total, and PCL-5 scores; the fourth row depicts results from the *post-hoc* adjustment controlling additionally for CTQ-abuse and CTQ-neglect scores; the fifth row depicts results from the *post-hoc* adjustment controlling additionally for a change in BDI-II scores between baseline and follow-up; the bottom row depicts results from the *post-hoc* adjustment controlling additionally for a change in STAI-total scores between baseline and follow-up. Colors reflect the z-statistic for the corresponding general linear (z-statistic>1.96; p<0.05 cluster-corrected for multiple comparisons). Volumetric brain maps are visualized on the surface for display purposes only; subcortical results in volume are shown with the left hemisphere on the left side of the image. SOMS:CD, Screening for Somatoform Symptoms-7 Subscale for Conversion Disorder; SSRI/SNRI, selective serotonin/norepinephrine reuptake inhibitor; FD, framewise displacement; BDI-II, Beck Depression Inventory-II; STAI-total, State Trait Anxiety Inventory-Total; PCL-5, PTSD Checklist-5; CTQ, Childhood Trauma Questionnaire.

**Supplementary Fig. 6.** Longitudinal changes in integration connectivity associated with physical symptom change (Δ PHQ-15 scores) between baseline and follow-up. (**A**) Regions with changes in integration that were significantly positively associated with physical symptom changes. Specifically, symptom improvement was associated with increased integration at follow-up compared to baseline in the left lateral occipital pole and occipital fusiform gyrus, while symptom worsening was associated with decreased integration in these regions. Colors in the map reflect the z-statistic for the primary general linear model adjusting for age, sex, SSRI/SNRI use, mean framewise displacement, elapsed time between baseline and follow-up sessions, and baseline PHQ-15 score (z-statistic>1.96; p<0.05 cluster-corrected for multiple comparisons). Statistical maps for additional post-hoc corrections are visualized in **Supplementary Fig. 6**. (**B**). Scatterplot of PHQ-15 change scores on the x-axis and mean integration change values averaged across significant voxels on the y-axis. While the analysis was conducted at the voxel-wise level, a Spearman correlation coefficient is included in the scatterplots to illustrate the strength of the association (rho=0.35). The shaded region reflects the 95% confidence interval. For visualization purposes, change scores are plotted with negative values on the right to reflect symptom improvement. (**C**) Bar chart summarizing between-network connectivity increases using all voxels in panel (A) as a visual network seed. Only networks with increased connections to the visual network seed at follow-up compared to baseline are shown, given that an increase in integration for these voxels was associated with symptom improvement. Connections with the frontoparietal, limbic, and default mode networks were higher at follow-up compared to baseline. F/U, follow-up; PHQ-15, Patient Health Questionnaire-15; VIS, visual network; FPN, frontoparietal network; LIM, limbic network; DMN, default mode network.

**Supplementary Fig. 7.** Results from analyses relating physical symptom change scores to changes in resting-state functional connectivity metrics (*left,* weighted-degree; *middle,* integration; *right*, segregation) between follow-up and baseline. The top row depicts results from the primary adjustment for age, sex, SSRI/SNRI use, mean framewise displacement (FD), elapsed time between baseline and follow-up sessions, and baseline PHQ-15 score; the second row depicts results from the *post-hoc* adjustment controlling additionally for FND phenotype (FND-seizure yes/no); the third row depicts results from the *post-hoc* adjustment controlling additionally for BDI-II, STAI-total, and PCL-5 scores; the fourth row depicts results from the *post-hoc* adjustment controlling additionally for CTQ-abuse and CTQ-neglect scores; the fifth row depicts results from the *post-hoc* adjustment controlling additionally for a change in BDI-II scores between baseline and follow-up; the bottom row depicts results from the *post-hoc* adjustment controlling additionally for a change in STAI-total scores between baseline and follow-up. Colors reflect the z-statistic for the corresponding general linear (z-statistic>1.96; p<0.05 cluster-corrected for multiple comparisons). Volumetric brain maps are visualized on the surface for display purposes only. PHQ-15, Patient Health Questionnaire-15; SSRI/SNRI, selective serotonin/norepinephrine reuptake inhibitor; FD, framewise displacement; BDI-II, Beck Depression Inventory-II; STAI-total, State Trait Anxiety Inventory-Total; PCL-5, PTSD Checklist-5; CTQ, Childhood Trauma Questionnaire.
