## Supplementary figures and images for "Functional Connectivity Predictors and Mechanisms of Symptom Change in Functional Neurological Disorder"

### Supplementary Figure 1

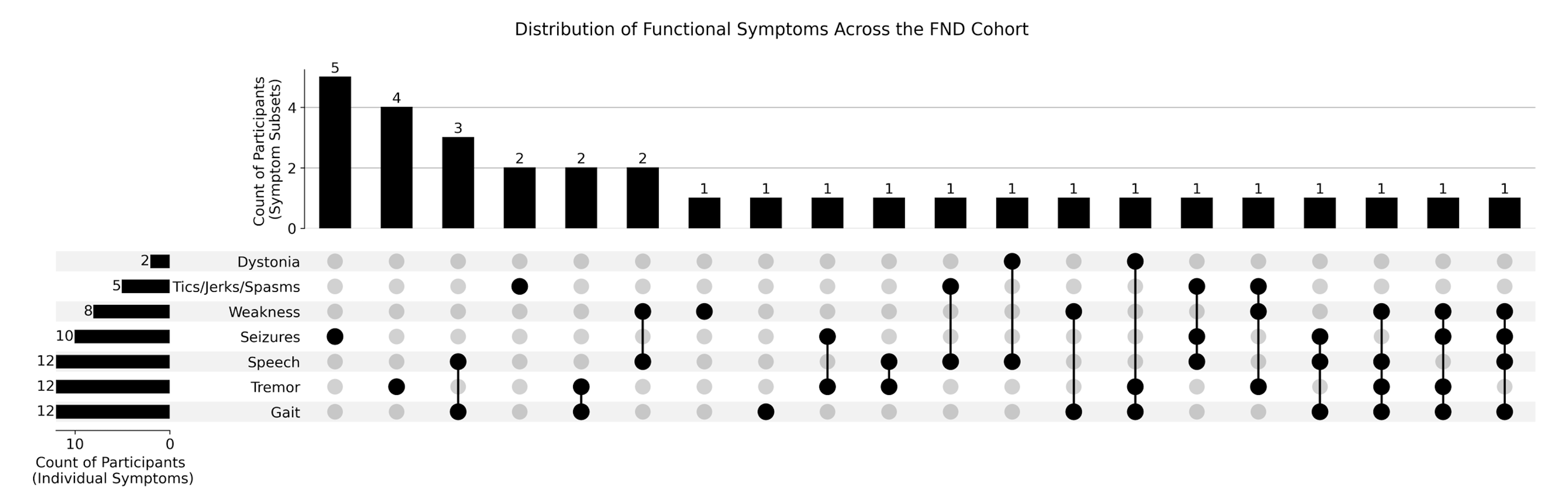

### Supplementary Figure 2

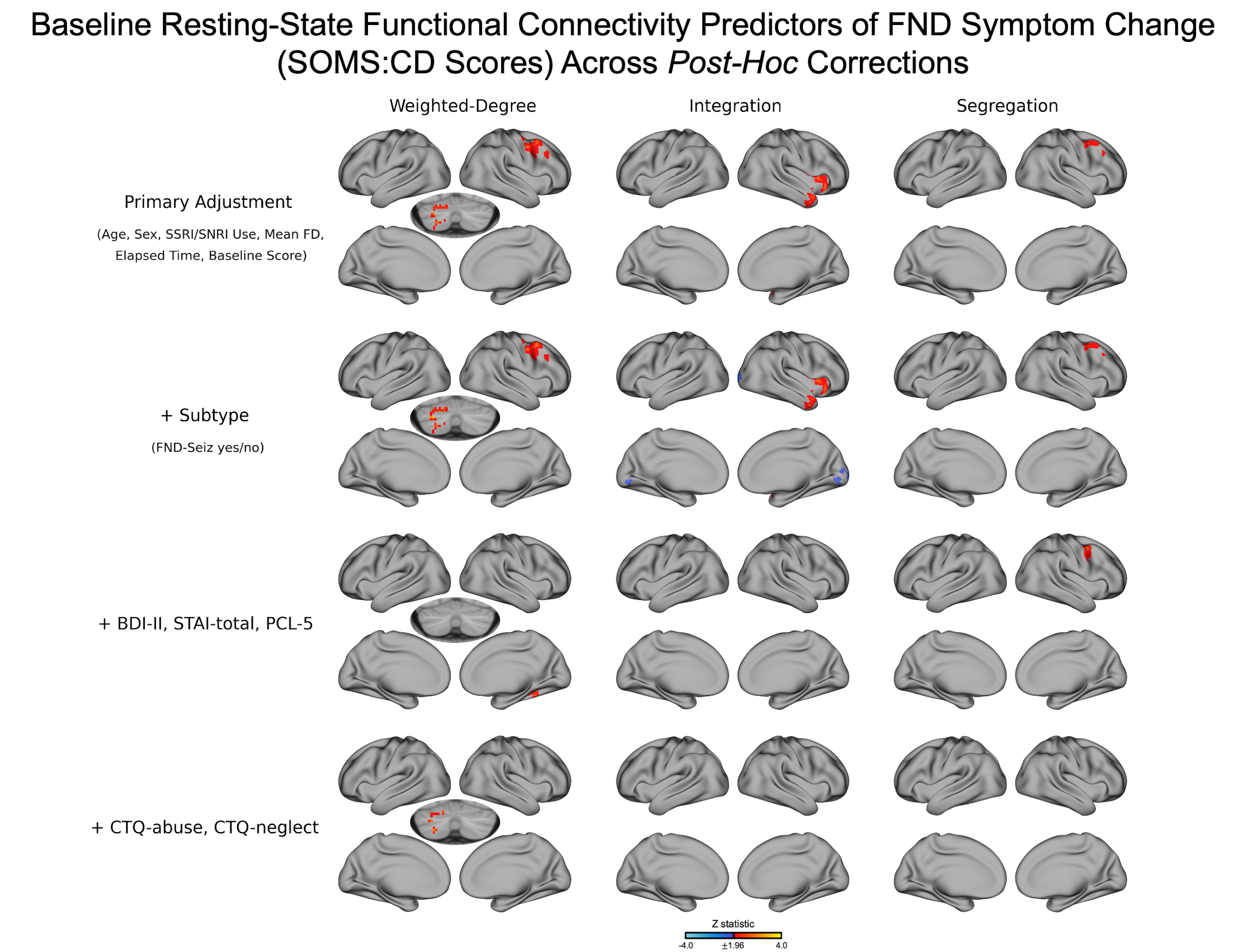

### Supplementary Figure 3

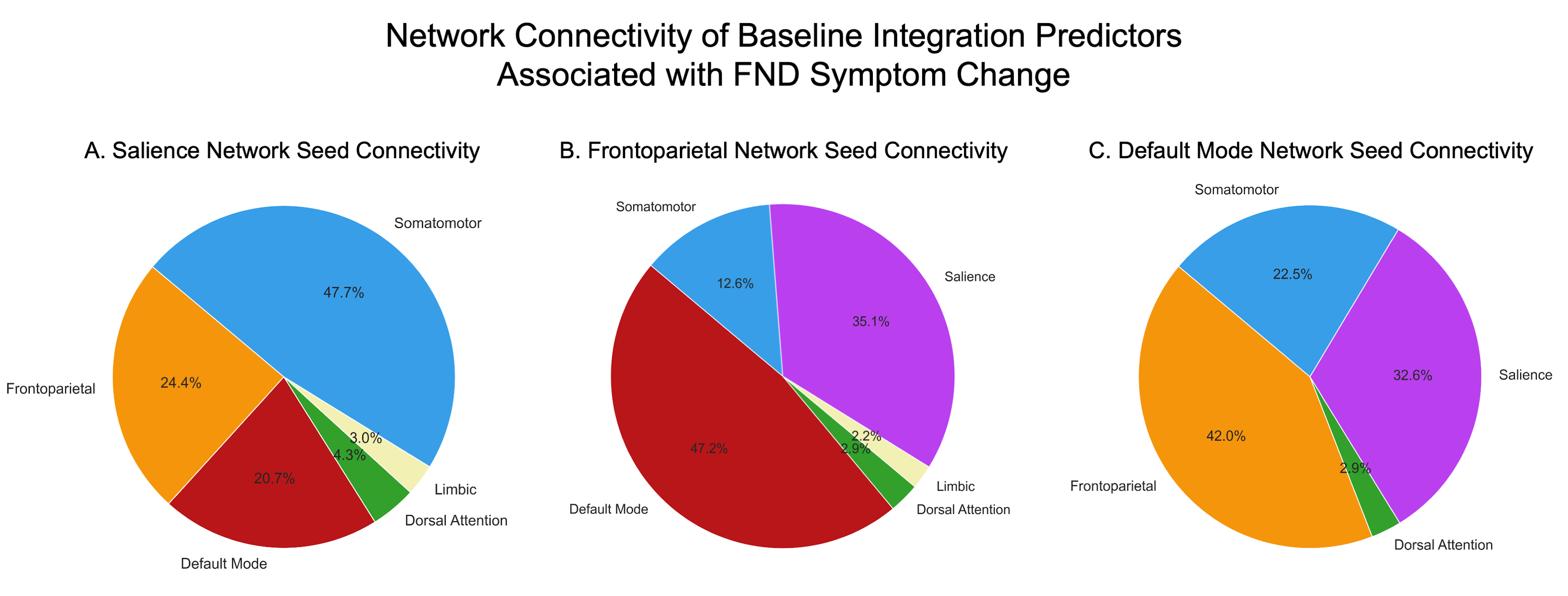

### Supplementary Figure 4

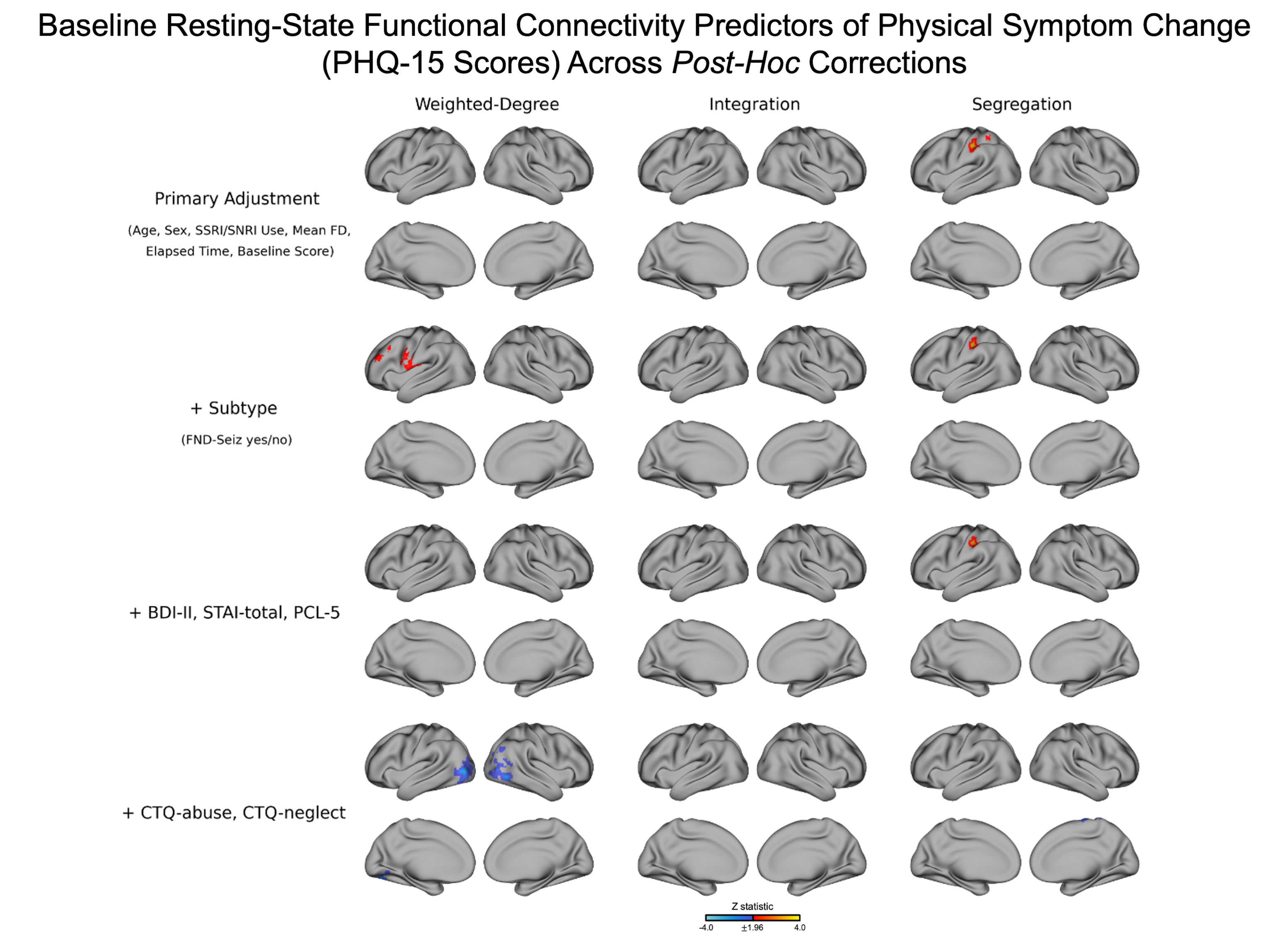

### Supplementary Figure 5

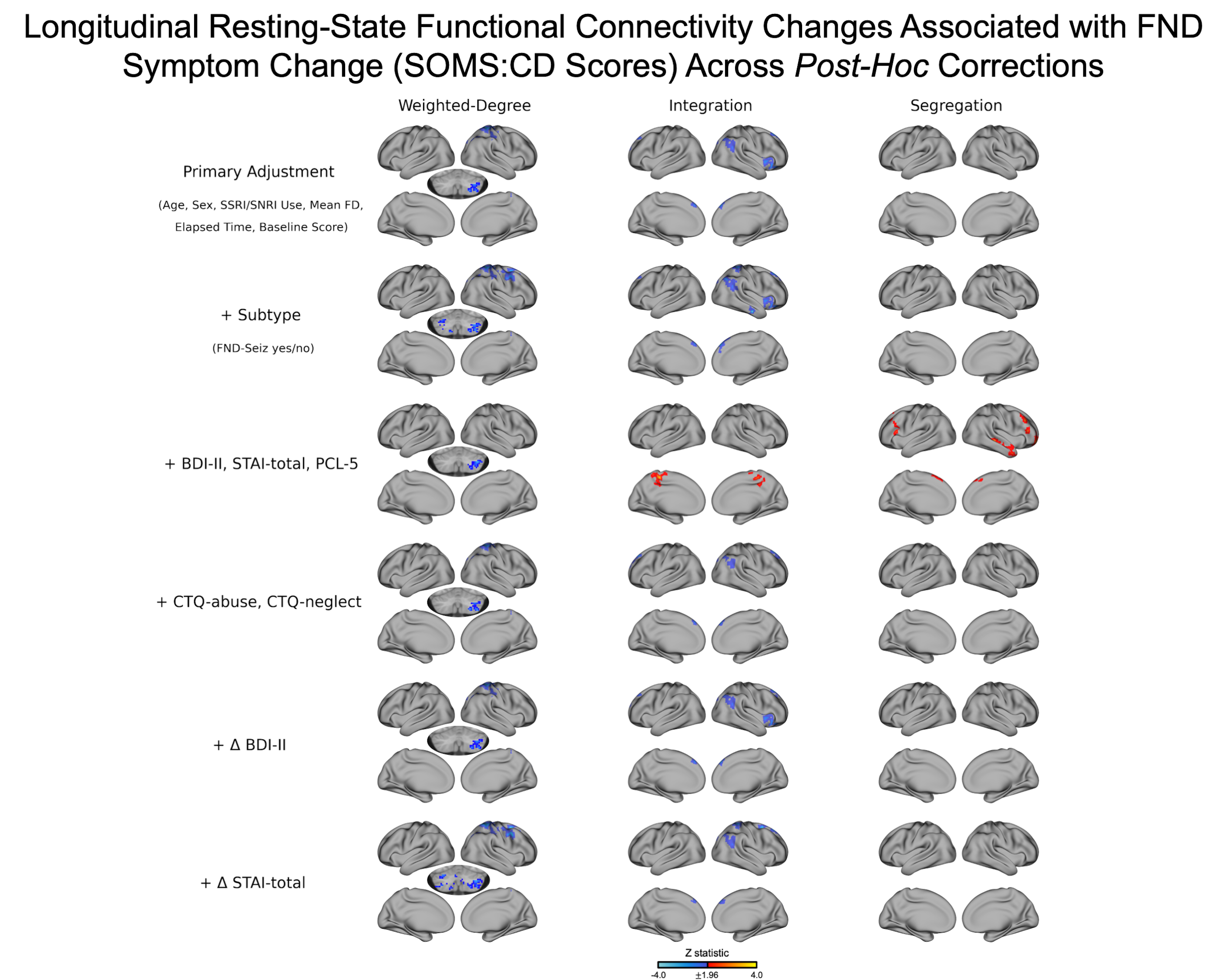

### Supplementary Figure 6

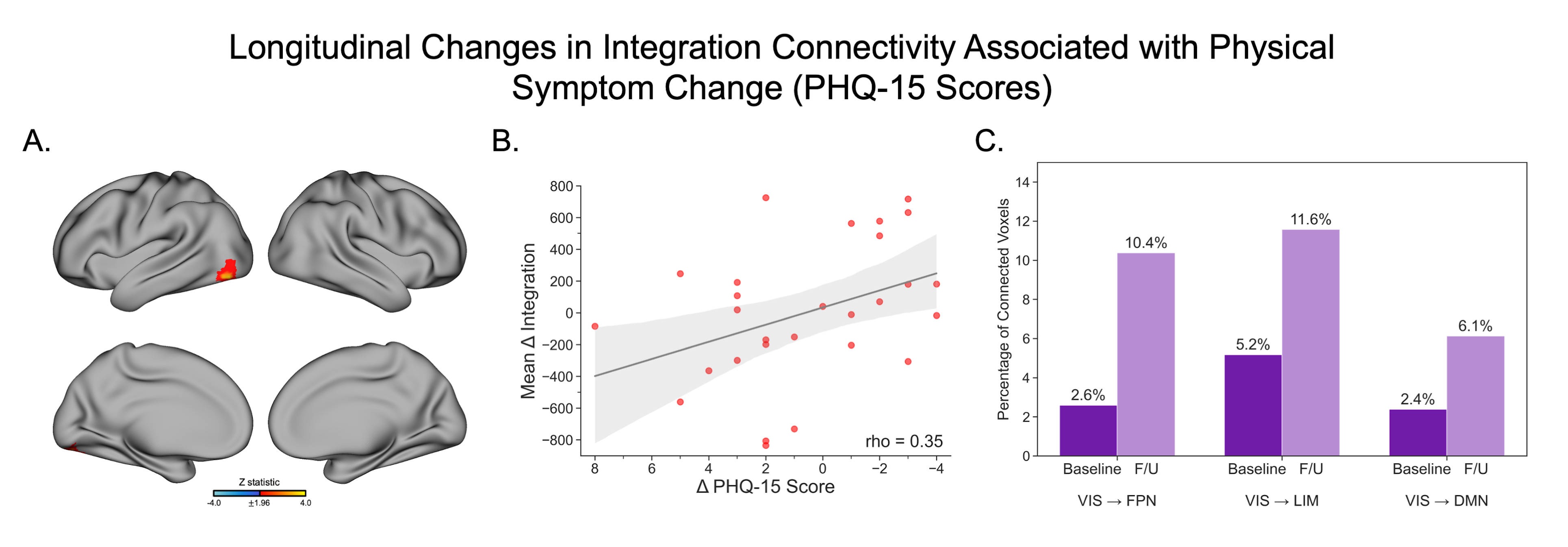

### Supplementary Figure 7

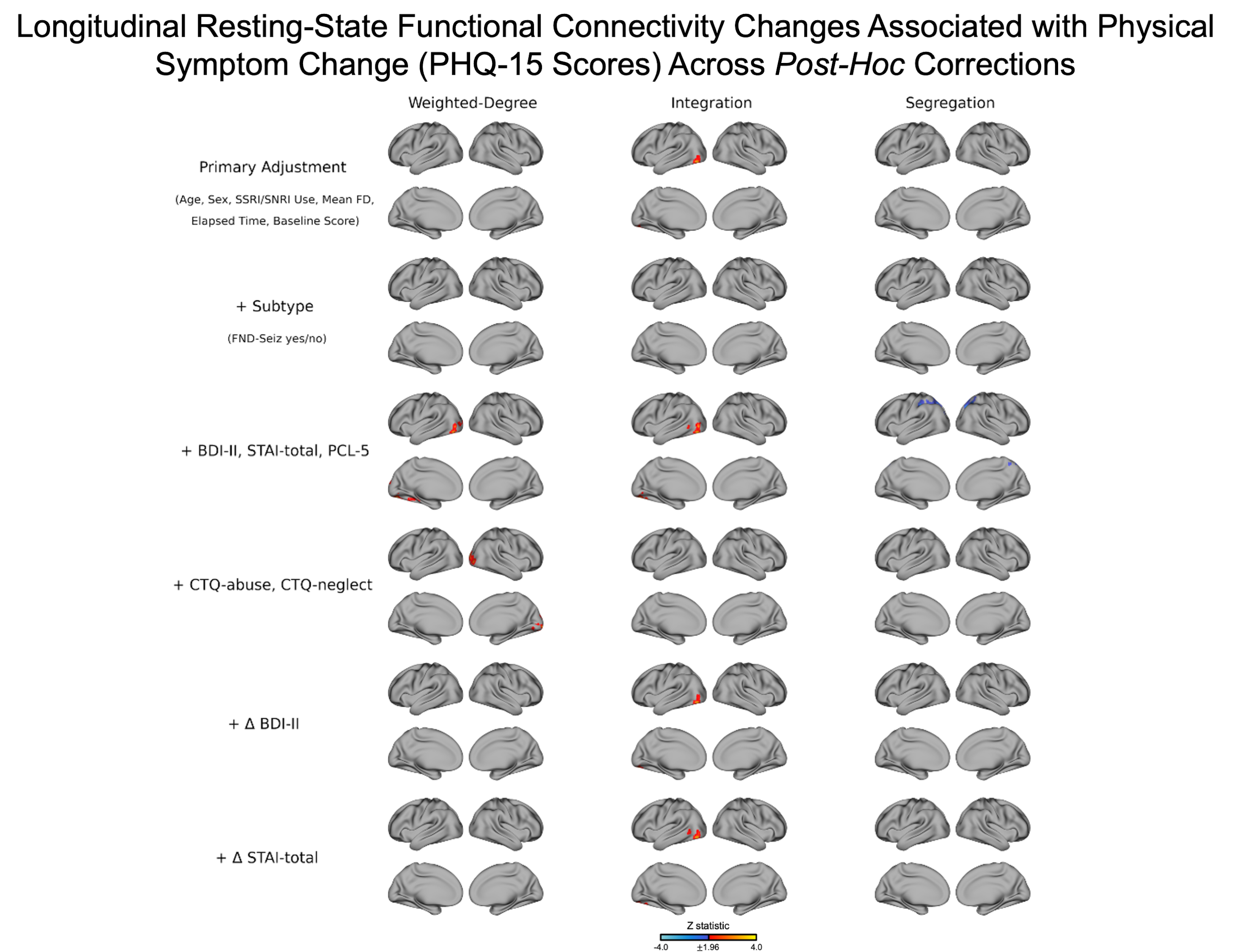
